# Scoping review and interpretation of Myofascial Pain/Fibromyalgia syndrome: an attempt to assemble a medical puzzle

**DOI:** 10.1101/2021.07.06.21260111

**Authors:** Shiloh Plaut

## Abstract

**Background:** Myofascial Pain Syndrome (MPS) is a common, overlooked, and underdiagnosed condition and has significant burden. MPS is often dismissed by clinicians while patients remain in pain for years. MPS can evolve into fibromyalgia, however, effective treatments for both are lacking due to absence of a clear mechanism. Many studies focus on central sensitization. Therefore, the purpose of this scoping review is to systematically search cross-disciplinary empirical studies of MPS, focusing on mechanical aspects, and suggest an organic mechanism explaining how it might evolve into fibromyalgia. Hopefully, it will advance our understanding of this disease.

**Methods:** Systematically searched multiple phrases in MEDLINE, EMBASE, COCHRANE, PEDro, and medRxiv, majority with no time limit. Inclusion/exclusion based on title and abstract, then full text inspection. Additional literature added on relevant side topics. Review follows PRISMA-ScR guidelines. PROSPERO yet to adapt registration for scoping reviews.

**Findings:** 799 records included. Fascia can adapt to various states by reversibly changing biomechanical and physical properties. Trigger points, tension, and pain are a hallmark of MPS. Myofibroblasts play a role in sustained myofascial tension. Tension can propagate in fascia, possibly supporting a tensegrity framework. Movement and mechanical interventions treat and prevent MPS, while living sedentarily predisposes to MPS and recurrence.

**Conclusions:** MPS can be seen as a pathological state of imbalance in a natural process; manifesting from the inherent properties of the fascia, triggered by a disrupted biomechanical interplay. MPS might evolve into fibromyalgia through deranged myofibroblast in connective tissue (“fascial armoring”). Movement is an underemployed requisite in modern lifestyle. Lifestyle is linked to pain and suffering. The mechanism of needling is suggested to be more mechanical than currently thought. A “global percutaneous needle fasciotomy” that respects tensegrity principles may treat MPS/fibromyalgia more effectively. “Functional-somatic syndromes” can be seen as one entity (myofibroblast-generated-tensegrity-tension), sharing a common rheuma-phycho-neurological mechanism.

## Introduction

Chronic pain is a major cause of morbidity and has a significant impact on quality of life [1]. Myofascial pain denotes a pain arising from muscle and fascia. Commonly known as “muscle knots” myofascial pain usually arises in trigger points (TrPs) or ‘tender spots’ [2–4]. TrPs are small and sensitive areas in a contracted muscle, that spontaneously or upon compression cause pain to a distant region, known as a referred pain zone [3,4]. Traditionally, “TrPs” are perceived as associated with MPS and differ from “tender points” mainly in that they radiate pain [2–4].

Myofascial pain syndrome (MPS) is an entity that still lacks a clear definition. Some define it as a regional pain disorder [2], others define it on the basis of tenderness and associated painful spots [3]. Nonetheless, TrPs and myofascial pain are a hallmark [2,4,5]. A fundamental difficulty arises when there is no clear definition, epidemiology, pathophysiology, or diagnosis of MPS. Having no accepted definition or criteria, clearly raises issues for both diagnosis and potential studies [6–8]. The purpose of this paper is to systematically search the empirical studies and components of MPS in an effort to assemble them into a suggested organic mechanism, explaining its pathophysiology and how it may evolve into fibromyalgia. Since much of the subject is still somewhat under dispute, textbooks and medical literature were added to establish a consensus on the elements of MPS.

In clinical practice MPS is often defined by multiple areas of musculoskeletal pain and tenderness associated with painful points [3]. Pain is deep and aching. It may arise after trauma, overuse or sedentarism [3]. A study found laborers who exercise heavily are less likely to develop manifestations of MPS than sedentary workers [3]. Palpation of a TrP can reproduce or accentuate the pain [4]. However, these findings are not unique to MPS: in a controlled study they were also present in “normal” subjects [3]. Literature estimates 45% of men and 54% of women in the general population have TrPs [2,9]. Estimates are 37 to 65 percent of the population have myofascial pain, which costs the united states $47 billion every year [4,10,11]. MPS is one the most frequently under-diagnosed, under-treated and misunderstood sources of the ubiquitous aches and pains of humankind [4,9]. However, even recently, MPS is considered by some to be fiction, or otherwise paired with psychosomatic disorders since it involves pain and has no clear pathophysiology [11–13].

Although MPS and fibromyalgia are defined as separate entities, the two may co-exist or considerably overlap [2,3]. They are both characterized by painful points, silent routine laboratory investigations and no systemic inflammation [2–4]. In some patients, regionally localized MPS may seem to evolve into fibromyalgia [3]. The puzzle of the mechanism of MPS and how it may develop into fibromyalgia still eludes our understanding. The marked dissociation between the estimated prevalence and burden of MPS, and the length of text on it in common medical textbooks, reflects this lack of understanding.

Studies of fibromyalgia reveal odd findings e.g., complete resolution after laparoscopic surgery, and strong overlap with other conditions (e.g., hypermobility syndrome). Many theories exist for fibromyalgia, the most accepted and studied seems to be central sensitization, but no single theory seems to explain a wide range of empirical evidence, and the pathophysiology and etiology are still not clear. Treatments are insufficient, meanwhile patients suffer. This scoping review focuses on the organic mechanical aspects of myofascial pain with the hope it might advance our understanding of MPS and fibromyalgia, highlight gaps in current knowledge, and stimulate research in a less explored field. Findings will be presented with the purpose of understanding the elements relevant to MPS and fascia, in order to assemble them and discuss a suggested mechanism for MPS and fibromyalgia. The discussion is divided into two parts: part 1 focuses on MPS and suggests a theoretical mechanism (“fascial armoring”), part 2 discusses empirical evidence in support of fibromyalgia as an entity driven by myofibroblast-generated-tensegrity-tension, as predicted by this model.

## Methods

A systematic search was held for multiple combinations of keywords in multiple databases. Keywords included : “fascia tension pain”, “trigger point satellite”, “fascia stiffness pain”, “Fascia Densification”, “trigger point densification movement”, “biotensegrity”, for all fields with no time limit; and for “Risk factors myofascial pain syndrome” only for title/abstract; Systematic reviews, meta-analyses and randomized controlled trials were searched in title or abstract for “myofascial pain syndrome”, “spinal mobilization sympathetic nervous system”, “sympathetic activity induced by pain” since 2015. The databases used were National Library of Medicine PUBMED (MEDLINE), COCHRANE, EMBASE, PEDro and medRxiv. Web of science was used to find items through a cited reference search. All searches were done between September 2020 and September 2021. Only items published in English were included. This work follows PRISMA guidelines for scoping reviews. PROSPERO yet to adapt registration for scoping reviews. A data charting form was used to abstract key data, as well as a summary document that was updated in an iterative process: that document evolved into this review. The systematic search yielded 979 items. After removal of duplicates (N=221), 758 items were screened by assessing both title and abstract. Items off topic, foreign language, and in journal ranked Q4 were excluded (N=364). For the remaining items (N=394) a full text determination of eligibility was then performed. Four items were excluded from the systematic search because they were off topic. Final number of items included in systematic search was N=390.

When side topics were encountered, literature was added to broaden scoping review via medical textbooks, www.uptodate.com and more studies from searching databases (n=409). These side topics were not searched systematically. An attempt was made to cite more than one source in such cases. Total review items: 799. Fig 1 shows the flowchart of the scoping review.

**Fig 1.**
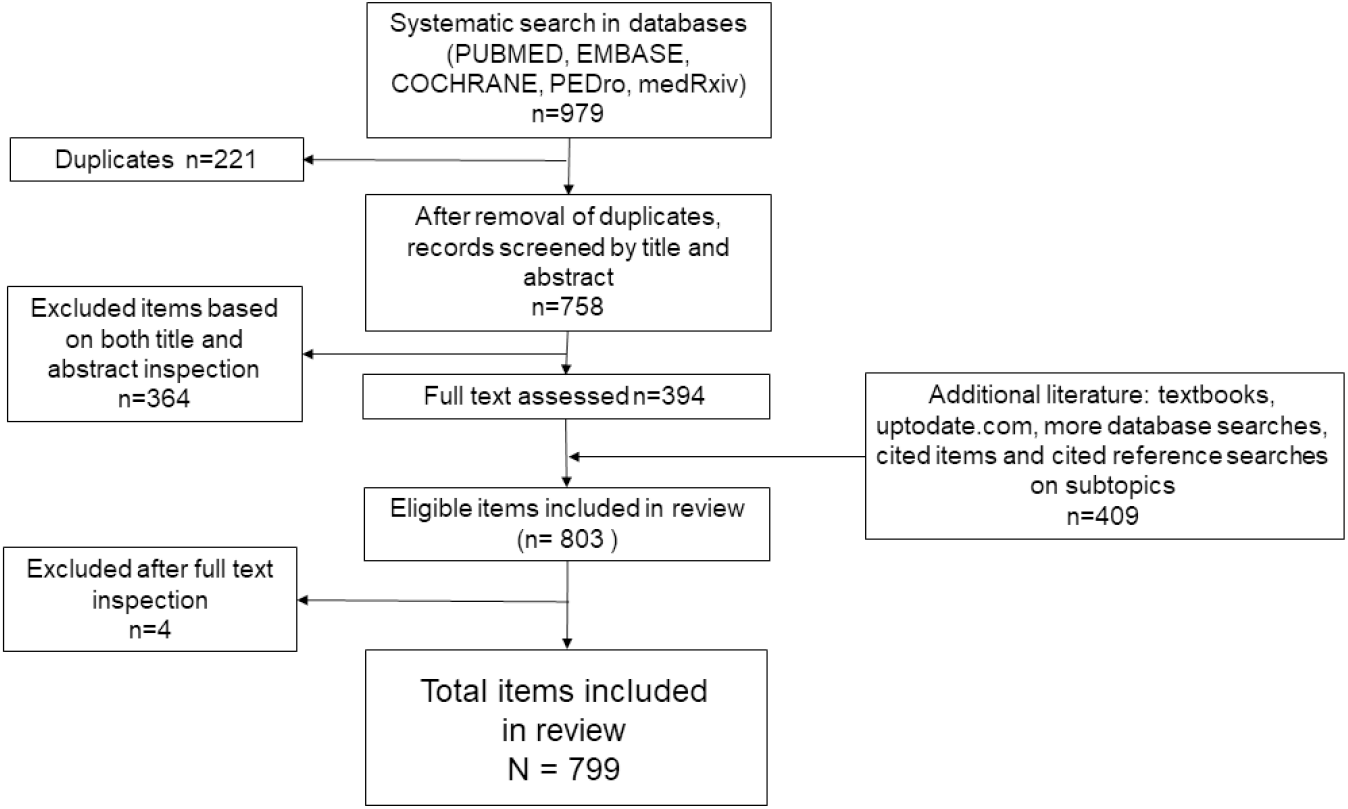
Flowchart of scoping review.

## Results

A total of 799 items were included. Numbers of items by recurrent themes were: “MPS and trigger points” (n=95), “properties of fascia” (n=47), “anatomy and movement” (n=43), “nervous system” (n=31), “biotensegrity” (n=26), “myofascial pain treatment” (n=209), “myofibroblasts” (n=93), “fibromyalgia” (n=93), “reviews” (n=33), and “other” e.g., somatic syndromes, stress, chronic pain, Dupuytren’s disease, plantar fasciitis (n=129). The systematic search included 127 clinical trials, 33 systematic review, and 19 meta-analyses. The findings are presented with the purpose of understanding the elements relevant to MPS and fascia, in order to assemble them and discuss a suggested mechanism of MPS and fibromyalgia. A microscopic view will set the stage for a macroscopic understanding.

### Properties of fascia

#### General

Fascia is an important yet often misunderstood tissue in medicine. Essentially, fascia is connective tissue composed of irregularly arranged collagen fibers, distinctly unlike the regularly arranged collagen fibers found in tendons, ligaments, or aponeurotic sheets [14]. When connective tissue collagen is pressed, it can chondrify [15]. Fascia pervasively extends from head to toe, it envelops and permeates muscles, bones, blood vessels, nerves, and viscera, composing various layers at different depths [16]. Fascia supports the human body in vital functions such as posture, movement, and homeostasis [14,16–18], as well as containing various sensory receptors for proprioception, nociception, and even hormones [18]. Nociception is influenced by the state of the fascia [18]. This means pain can arise from changes in the connective tissue [18,19]. The interconnectedness of fascia and its ability to transmit force are at the basis of its functions [18]. Fascia is continuous from the trunk across the upper and lower limbs and hence has the potential to influence range of motion [18]. Fascia seems to connect the distant hip and the ankles, not only anatomically, but also mechanically, supporting the concept of a myofascial continuity [18].

#### Biophysical and mechanical functions

Fascia has properties that enable it to reversibly change its stiffness and decrease the forces of tension experienced by it. This is a mechanical ‘stress relaxation’ resulting from the viscoelasticity of fascia that expresses high or low stiffness, depending on the rate of loading. Some experimental tests demonstrate a 90% of stress relaxation achieved in about four minutes, afterwards the stress relaxation curve is near linear horizontal [20–23]. Other studies observe increase in stiffness when ligaments are successively stretched, i.e., strains produced by successive and identical loads decrease. They recover to baseline after a rest period. Isometrically maintained stress resulted in gradual tightening of the tissue [14].

Blood vessels and nerves are interspersed in fascia, therefore, involvement of these structures due to fascial changes is commonly seen [14,19]. It is noted a tender point in the gluteus medius can refer pain down the leg and mimic sciatica [3]. Often the patient is aware of numbness or paresthesia rather than pain [9]. Pain is the result of a microenvironment around the nerve composed of connective tissues (e.g., deep fascia, epineurium). Tissue modifications can be translated into change in nerve mobility, with a consequent decrease in the independency of the nerve from its surroundings [24,25]. Altered fibrous tissue around nerves can lead to entrapments and lesions [24,25]. Circulation and perfusion can also be compromised. For example, dysfunction of the posterior layer of thoracolumbar fascia has been reported as a “chronic compartment syndrome” of the paraspinal muscles [26].

#### Hyaluronic acid

Hyaluronic acid (HA) has a key role for fascia. It is the major glycosaminoglycan of the extracellular matrix (ECM) and a major constituent of connective tissue [27]. Its concentration, as well as the temperature and other physical parameters, determine the physical properties of the ECM [27–30]. In fascia, HA is present within the sublayers and facilitates normal free sliding [27]. Infra-red spectroscopy of viscous HA indicates water molecules can be arranged tetrahedrally almost like as in ice [29]. HA interchain interactions are reversible, disaggregation occurring with an increase in pH and temperature; a gel-like to fluid-like transition occurs at 35-40 °C [29]. HA solutions can express high viscosity and non-Newtonian flow properties [28], and HA’s osmotic activity is relatively high; Its stiffness occurs in part due to its long chains forming an entangled large volume network [29,30]. Once viscosity increases, HA can no longer function as an effective lubricant; this increases resistance in layers with abnormal sliding [27].

### Fascia and movement

Fascia acts as a key player for generating movement [16]. Although often a dropped prefix, fascia is part of the (fascio)musculoskeletal system. It is suggested to be able to transmit tension and affect other muscles, reflecting the direction of force vectors, and play a role in properly coordinated movements of the body [31].

#### Sedentarism

Non-versatile movement patterns and sedentarism are an important lifestyle component for fascial changes and pain [3]. A meta-analysis found that low work task variation is a risk factor for non-specific neck pain [32]. Immobilization of a limb or body segment can lead to pathological changes in the connective tissue. Studies of immobilization suggest an increase in collagen and myofibroblast density as early as one week after immobilization [33]. After about 4 weeks of immobility in the shortened position the collagen fibrils become arranged abnormally [34]. Changes start in the perimysium and after longer periods of immobility the endomysium becomes involved too [34]. Regular physical activity is recommended for treatment of chronic pain and its effectiveness has been established in clinical trials for people with a variety of pain conditions [35]. Proper posture and resting positions are important in preventing muscle tension [3]. Pain often recurs unless appropriate exercises are prescribed [5].

#### Tensegrity

The fact that fascia can transmit tension to a distance is a basis for a “bio-tensegrity” framework [36,37]. Bio-tensegrity applies principles of tensegrity to our understanding of human movement. Tensegrity is an architectural principle. According to this, structures (or tensegrity systems) are stabilized by continuous tension with discontinuous compression, and function as one structure [37].

When the tension of fascia increases, the connective tissue can distribute the forces throughout the surrounding areas, propagating along the myofascial system [14,18]. The forces passively imposed in a muscle by stretching are distributed over the tissue as a whole by means of the intramuscular connective tissue [34]. According to Wilke et al. [38] fascia links the skeletal muscles, forming a body-wide network of multidirectional myofascial continuity. Cadaveric studies suggest a clinically relevant myofascial force transmission to neighboring structures in the course of muscle-fascia chains (e.g., between leg and trunk) [38]. A study suggests manual traction to the biceps femoris tendon results in displacement of the deep lamina of the thoracolumbar fascia up to level L5–S1 [14]. Acute bout of stretching of the lower limbs increases the maximal range of motion of the distant upper limbs and vice versa [18]. Recent studies indicate that tightness of the gastrocnemius and hamstrings are associated with plantar fasciitis [39]. Since most skeletal muscles of the human body are directly linked by connective tissue, symptoms may develop in areas distant from the locus of dysfunction [40].

Acceptance of the concept that a fascial tensegrity system connects the whole body is not necessary for a scientific discussion of tension spreading to other areas of muscle groups or structures. Some studies indicate that a tensegrity model might not be a true representation of the whole fasciomusculoskeletal system, and some evidence supports tensional propagation only to certain areas [41].

### Myofascial Pain Syndrome

There are several phenomena that accompany MPS including: trigger points (TrPs), active loci, taut bands, satellites, and the local twitch response.

Trigger points - Changes in fascia are associated with pain in TrPs and tender spots. Many patients with active (i.e., painful) TrPs have other areas with the same electrophysiologic findings that are not symptomatic, called latent TrPs. Latent TrPs seem to only cause pain upon palpation, while active TrPs cause pain and symptoms both at the site palpated and referred elsewhere [2,4,9]. The arising of pain is attributed to those palpable areas in the connective tissue, which seem to activate nociception [2,4].

Theories of the origin of TrPs emphasize the chronic contraction associated with them or their tendency of appearing at the muscle spindle [2,9]. Others suggest TrPs represent hyperactive end-plate regions [7]. More theories that either deny the existence of MPS or believe it represents a focal dystonia, microtrauma, or are of central nervous system origin, are also hypothesized [2,7]. Critics of MPS argue that the findings seen at TrPs are simply variants of a normal physiologic finding [7]. There is reason to suspect this is true: about 50% of adults have active or latent TrPs [2,9], and infants have been observed with point tenderness of the rectus abdominis muscle and colic. These both were relieved by sweeping a stream of vapocoolant, which inactivates myofascial TrPs, over the muscle [5].

TrPs are extremely common. Among 224 non-specific neck pain patients, TrPs were found in all of them [9]. In studies in pain clinics, 74-85% of those presented to a clinic had TrPs the primary cause of their pain [9]. TrPs are one of the most frequent causes of neck pain and back pain [42]. The severity of symptoms caused by TrPs ranges from the agonizing, incapacitating pain caused by extremely active TrPs to the painless restriction of movement and distorted posture caused by latent TrPs [5]. The influence of latent TrPs on physical function is commonly overlooked [5]. Patients who had other kinds of severe pain, such as that caused by a heart attack, broken bones, or renal colic, say that the myofascial pain can be just as severe [5]. Some suggest TrPs are present in up to 85% of individuals with colorectal, urological and gynecological pelvic pain syndromes, and can be responsible for many of the symptoms related to these syndromes [43]. It was suggested that TrPs or myofascial tension, not exerted by external forces, can apply enough force to cause various abnormalities, including compression and entrapment of anatomical structures [44], decrease joint range of movement [7,16], affect visceral organs [9,45,46], lead to musculoskeletal abnormalities [47], and alter blood and lymph flow [48].

Active loci - TrPs show electrical abnormalities called ‘active loci’. This is a small region in a muscle that exhibits spontaneous electrical activity (SEA), often characterized as endplate noise measured on electromyography (EMG), leading to chronic contractions [9]. Although some literature does not regard TrPs as a discriminating factor for MPS, the ‘Active Locus’ seems to be a consensus [7].

Taut Band - A taut band is thought to be composed of several TrPs and shows excessive endplate potential activity [9]. Sustained abnormal activation of acetylcholine (Ach) is hypothesized to create an “Energy Crisis” component [9].

TrP Satellite - TrP satellites are another disputed aspect of MPS. Some literature states TrPs themselves can induce motor activity (referred spasm) in other muscles [49]. Illustrated by an example where pressure on a TrP in the right soleus muscle induced a strong spasm response in the right lumbar paraspinal muscles. Pressure applied to a TrP in the long head of the triceps brachii muscle indued a strong motor unit response in the upper trapezius muscle only during the 20 seconds that pressure was being applied. This response failed to be reproduced following inactivation of the triceps TrP. It is known as a ‘satellite’ phenomenon [9,49]. Moreover, it seems TrPs can spread, since an active TrP in one muscle can induce an active satellite TrP in another muscle [9,49].

Various hypotheses have been suggested to explain the mechanism of MPS, among them are trigger points, non-muscular sensitization of the nervi nervorum, central nervous system, and several other theories [2,7,9,50]. Some relate the muscle contractions in TrPs to a myotatic (stretch) reflex evoked by fascial tension [7,9].

### Myofibroblasts

Fascia itself is able to actively contract. Tensional alterations are caused by contractile cells [51]. Myofibroblasts are present in some developing or normal adult tissues, altering tissue tension [30,51]. Although most tissues exist under a mechanical tension, the same is not necessarily true of their resident cells; These are protected from the relatively massive external loads by the mechanical properties of the surrounding matrix [51]. In engineering terms, this is called ‘stress-shielding’, occurring due to the matrix material that they deposit and remodel [51].

Normal fibroblasts are highly sensitive to the physical stimuli. Tomasek et al. [51] show certain changes in tissue rigidity, strain, and shear forces are mechanical cues sensed by fibroblasts that lead them to trans-differentiate into another cell phenotype [51]. In other words, the transition from fibroblasts to myofibroblasts is influenced by mechanical stress. If subjected to mechanical tension, fibroblasts will differentiate into proto-myofibroblasts, which contain cytoplasmic actin stress fibres that terminate in fibronexus adhesion complexes [51,52]. The adhesion complex bridges the myofibroblast’s internal cytoskeleton and integrins with the ECM fibronectin fibrils. This enables proto/myofibroblasts to generate contractile force in the nearby ECM by traction. The force inside the ECM is maintained over time and reinforced further by remodeling and collagen deposition [51]. Functionally, this provides a mechano-transduction system, so that the force that is generated can be transmitted to the surrounding ECM [51]. Increased expression of alpha smooth muscle actin (α-SMA) is directly correlated with increased force generation by myofibroblasts in a positive feedback regulation [51]. A vicious cycle is suggested, in which tension facilitates TGF-β1 signaling, which induces α-SMA expression. In turn, this increases development of more tension, which upregulates TGF-β and α-SMA repeatedly [51]. In short, myofibroblasts generate the mechanical conditions that enhance their contractility in a detrimental loop [51]. In contrast to the reversible short-lived contraction of striated and smooth muscles, myofibroblast contractility is different: with ECM synthesis and degradation (i.e., remodeling) they lead to irreversible and long contractures in a process that can be maintained for long periods of time [51]. It is thought that myofibroblasts use a lockstep or ‘slip and ratchet’ mechanism, in incremental and cyclic contractile events using Rho-kinase system, and aside the myosin light chain kinase (MLCK) system [51]. Once achieved, contracture shortening does not require the continuing action of myofibroblasts [51]. The visible appearance of continuous tension in pathological contractures is the consequence of contraction and remodeling [51]. Myofibroblasts might transmit considerably high forces [51].

It is also possible that myofibroblasts can couple their activity directly to other cells like myocytes via gap junctions and act as a unite [51,53]. Interestingly, if the collagen lattice is released from its points of attachment such that stress in the matrix is lost, the cells rapidly undergo isotonic contraction and subsequently lose their stress fibres and fibronexus adhesion complexes [51].

### Treatments for MPS

Several pharmacological and non-pharmacological treatments are suggested in literature.

Non-pharmacological [4,9,11,54–57]: including manipulative therapy, vibration therapy, exercise, etc. Since it is shown that needle insertion reduces pain and SEA on EMG [58], this suggests a therapeutic role for mechanical needling.

Pharmacological [4,9–11,54]: including non-steroidal anti-inflammatory drugs, opioids, topical creams, and TrP injections.

## Discussion

The discussion includes two parts. Part 1 discusses myofascial pain syndrome (MPS) and presents a suggested theoretical model which is based on the scoping and systematic literature search. Part 2 is based on part 1 and discusses implications derived from the model and further empirical findings in support of the main hypothesis predicted by this model.

### Part 1 – A theoretical model for MPS

Synthesis of the data gathered in this review leads the author to the following

#### Etiological considerations

There are three aspects of fascial induced pain to consider, from the standpoints of physics, biomechanics, and cellular biology. These seem relevant to fascial tension, TrPs, and MPS.

The standpoint of physics relates to energy. Thermodynamically, the human body can be seen as an open system of energy, and so can the fascial system. Although energy and entropy are modulated by various factors, by nature, entropy of this system will spontaneously tend to increase. To tense the fascia energy needs to be expended. Once fascia is tense the energy can remain as potential elastic energy or change due to other modifications, for example molecular rearrangement. Hyaluronic acid (HA) has a molecular structure that enables it to have a high degree of entropy, like an entropic “sponge”. When entropy increases it generates osmotic pressure and diffusion of fluids and other processes. Thus, when entropy of HA increases, osmotic pressure increases, and stress decays. If stimulus is not removed after this acute stressing, fascia will remain in the failure part of the stress relaxation curve and will be remodeled in this state. Although entropy goes together with osmosis and stress decay, each oscillation between different energy states will likely cause a loss of energy being a non-ideal system, for example due to friction. Fascia has hysteresis and is not a perfect “energetic spring” to oscillate between high-tension and low-tension states, i.e., it dissipates energy.

Secondly, from a biomechanical standpoint, the literature review shows that (i) simple stress decay and recovery from it is a reversible process due to elasticity of fascia; (ii) trigger points (TrPs) and myofibroblasts are found in normal individuals; TrPs are seen even in childhood and infancy, and in animals [58]; (iii) fascia continuously remodels itself and HA is continuously synthesized and excreted; (iv) a sedentary lifestyle and repetitive overuse of muscles, with low variability of movements, all lead to fascial changes/fibrosis and possibly MPS; (v) exercise and proper movement relieve and prevent MPS; (vi) MPS seems to recur unless appropriate exercises are prescribed.

These suggest an underlying ongoing interplay between movement and sedentarism. Since tensional changes are reversible in fascia of normal individuals and depend on the mechanical state of the human body, this indicates fascia is meant to withstand and continuously undergo dynamic changes according to varying dynamic mechanical states. Since immobility often leads to collagen alterations, HA changes, myofibroblasts, and pain, we can see MPS as a natural consequence of the sedentary lifestyle. Resulting from the evolutionary “price” biology paid in a trade-off with chemistry when trying to “come up” with a tissue as effective, pliable, compliant and strong as the fascia. This “bargain” served us well to function effectively as an organism dominated by continuous movement. It seems that by living sedentarily, the natural disadvantages of the properties of fascia (chondrifying, HA aggregation, failure to decay, and fibrosis) manifest pathologically. Stiffness and lack of mobility of fascia has implications beyond a patient being unable to move adequately, it can affect the behavior of all cells interacting with the connective tissue matrix [16]. If indeed nature designed a way to relax fascia during movements and reverse these changes it might happen through (i) dissipation of fascial energy as mentioned above; (ii) mechanical effect of movement that ruptures fascial fibers or shakes off fibers, which might explain why studies find extra-corporeal shock wave therapy to be effective for MPS (e.g., Uritis et al. [54]); (iii) friction between sliding layers of fascia elevating the local tissue temperature, or “warming up” myofascial tissue. Literature suggests the three-dimensional supramolecular assembly of HA breaks down progressively at 35-40 degrees Celsius [29]. The more viscous is HA, the more friction will act to counter viscosity; (iv) increasing clearance of HA due to increased lymph flow. Higher concentrations of HA lead to its more gel-like state, affecting the properties of the matrix; (v) pandiculation; (vi) self-aware “palpation” as a part of one’s lifestyle.

The third aspect to consider is a cellular one: induction of myofibroblast by tension and by sedentarism [33,51]. Chronic tension is exacerbated with more tension generated by myofibroblast smooth muscle actin fiber contractions. This makes it increasingly more difficult to maintain a relaxation. Tension is converted to HA aggregation and entropy so long as compensation along the stress relaxation curve can allow for it. Increase in HA concentration is expected to keep HA in the denser gel-like state [29,59], which would plausibly be perceived by the individual as bodily stiffness. If the values of tension reached at the stress relaxation curve plateau tend be above the value of the threshold for myofibroblast differentiation, it will be a major driver for MPS (values may vary between individuals). Biologically, the reinforcing of fascial cells with actin fibers may be the way of the body to express support of the repetitive muscular effort. Accumulation of many foci of stretching along a sensory or sympathetic nerve, due to extracellular fibers pulled from multiple directions by proto/myofibroblasts, might cause peripheral nerves to be hyperirritable chronically. This may explain why a study finds the number of active and latent TrPs is significantly and negatively associated with pressure pain thresholds (Do et al. [12]). A chronic peripheral pain could contribute to a central sensitization and to changes in spinal cord pathways, though, as stated, the central mechanisms are not the focus of this review. These three dimensions seem to be relevant to fascial tension, TrPs and the etiology of MPS.

#### Pathophysiological considerations

Three elements combined might help explain MPS. These are: (i) tensegrity, (ii) TrPs, and (iii) myofibroblast contraction and stress shielding.

Manifestation of MPS is seen in TrPs. Latent TrPs may be points that have previously nociceptively sensed tension (i.e., active TrPs), due to sedentary behavior or a ‘satellite’, and then were mechanically induced to stress shield themselves via myofibroblasts. The stress shielding does not necessarily eliminate tension (and pain) completely, thus preserving some mechanical drive for further proto/myofibroblast activity. Moreover, new myofibroblasts generate alpha smooth muscle actin (α-SMA) and contractions themselves [51]. Over time, especially if mechanical stimuli continue, fascia remains more towards the failure part of the stress relaxation curve, and cells have time for more stress shielding and matrix remodeling. Then, latent TrPs (via myofibroblasts) start to exert their own tension. This reinforces the abnormality. However, that tension would be more diffuse as it spreads along an intricate web of fibers (i.e., stress shield), inducing more tensions, more force gradients in fascia, more foci of stretch or entrapments of nerves, contractures, and other painful TrPs further away. If mechanical tension is sufficiently reduced by shielding or by other means, myofibroblast will dedifferentiate or undergo apoptosis [51], leaving behind a remodeled dysfunctional fascia. Multiple iterations of the contractile cycle result in incremental and irreversible tissue contracture [60]. TrPs appear on ultrasound as focal, hypoechoic regions and with reduced vibration amplitude, indicating increased stiffness [12].

These dynamics may be the basis for the unexplained “metastasis” of TrPs and satellites that Quinter et al. [7] refer to. When sedentary behavior continues, it feeds more tension down the cascade. The clinical manifestation would be determined not only by the amount of myofibroblasts, but by the interconnectedness of their adhesion complexes, gap junctions, and fiber-cellular networks as well. Studies suggest myo/fibroblasts form a cellular network and can exhibit coordinated calcium oscillations and actively respond to mechanical stimuli [61–63]. Studies suggest myo/fibroblasts have stretch activated calcium channels, and that intracellular calcium and myofibroblast contractility is mechanistically linked [60,61]. Langevin et al. [62] find evidence that leads them to a conclusion that soft tissue fibroblasts form an extensively interconnected cellular network, suggesting they may have important and so far unsuspected integrative functions at the level of the whole body.

Activating a latent TrP is hypothesized to involve interference with the stress shield or stretch-activating myofibroblasts to rapidly contract, thus exposing the area to higher tension and initiate nociception. Eliminating a TrP seems to induce a local twitch response, which possibly signifies the neuron’s or muscle’s calcium driven response to a recovery from an active locus or shielded tension. Alternatively, it may be on-off “flickering” of the myotatic reflex due to nearby myofibroblast rapid MLCK contractions and their waves of calcium flux. Active loci may be a multifactorial phenomenon. For example, stress shielding (with or without gap junctions) superimposed on sympathetic activity, i.e., a myotatic reflex on top of sympathetic drive. It would seem plausible to expect one to “learn to relax” or “learn to cognitively override the sympathetic aspect by focusing attention”, but not to willfully override a reflex response. The fact that muscle spindles are surrounded by a capsule of connective tissue, an area that sometimes might be palpated as a nodule (i.e., TrP) suggests that if there are microscopic tensions, they might possess a sensitivity to generate more tension at the spindle (via proto/myofibroblasts positive feedback), thus locking the stretch reflex to a certain degree. Locking the reflex would “turn on” an active locus in the associated muscle. To complicate the above, if electrical coupling of this structure occurs with an end plate (or non-intrafuseal fibers), via myofibroblasts, it might perpetuate an “arrhythmic” picture on EMG, with background noise, and random spikes upon myofibroblast contractions. Mechanical disruption of this structure e.g., manipulative therapy, should unlock this pathology, at least partially. This might be one of the reasons for different degrees of body relaxation and the inability of some to experience it. If neurotransmitter molecules (or acetate) leak outside the synaptic cleft they might induce adjacent fibroblasts to transdifferentiate, which might explain why muscle overuse is associated with “TrPs” (“TrPs” can be found in peri muscular and myofascial tissue [4]).

“TrPs” and “tender spots” are therefore suggested here to be basically the same (driven by myofibroblasts), but different in their location and fascial layer. This notion is made under the now common presupposition that the main difference between the definition of “TrPs” and the definition of “tender spots” is radiation of pain. If a large nerve is affected by a nearby population of myofibroblasts that contract upon mechanical stimuli, it could radiate pain. If a large nerve is not affected by them, it might not radiate pain. If enough stress shielding and armoring has been created, nerves will be mechanically shielded from an external force applied to the tensegrity (i.e., palpation). Snapping palpation held longer would elicit more of a response (pp. 82 Travel, Simons & Simons [9]). Thus, the terms “TrPs” and “tender spots” can be used interchangeably throughout this discussion since both are suggested to be phenomenological variations of the same underlying biology: myofibroblast-generated-tensegrity-tension.

Notwithstanding, other factors are not excluded. Factors associated with the central nervous system could also be implicated in the genesis of MPS, as well as genetics, environmental factors, psychosocial aspects, chronic sympathetic “freeze” reaction to everyday psychological stress, etc. Based on this discussion, we might expect factors affecting fascial myofibroblasts to have potential to cause tension, pain, or even lead to MPS. For example, diet is believed to play a role in MPS [9], and studies suggest myofibroblasts are promoted by certain diets and by pesticides/herbicides [64–67]. Similarly, the molecule CCN2/CTGF is suggested to play a role in development of pain due to overuse [68], and CCN2/CTGF is shown to be important in myofibroblast α-SMA synthesis [69]. Applying this sort of logic, we may find more factors linking myofibroblasts to myofascial pain [70–73]

It is unclear where the tipping point between a status quo of movement-sedentarism and a deranged myofibroblast activity is. It is likely multifactorial, but lifestyle seems to be a major factor. A mechanical stimulus causes differentiation to myofibroblast [51,74], some studies suggest a threshold of ∼20-24 kPa [75]. Therefore, mechanical forces might be one of other possible factors contributing to MPS and myofibroblast induction. For example: sedentarism and muscle overuse [3,9,76–78], infection or inflammation [4,7], and trauma/fracture or immobilizing a casted limb [4,9,74,79,80]. Since MPS is so prevalent, also common overlooked factors in a lifestyle that is not evolution-oriented may be suspected, e.g., medications that are shown to both induce myofibroblasts and increase risk of myofascial pain [72,73].

#### Symptomatologic considerations

TrPs and myofascial pain, particularly when severe, can cause various symptoms that are not limited to the fascial-muscular-skeletal system. It is suggested that the sympathetic nervous system (SNS) causes TrPs [4]. Reciprocally, pain and TrPs can activate the SNS [2,4]. This activation may actually be due to coactivation of larger diameter mechano-sensitive afferents rather than nociceptive afferents [81,82]. If the SNS is activated by paraspinal mechanical stimuli, then maybe it can be activated by muscular tension alone. If so, this may serve as a slippery slope for further TrPs, active loci and MPS. Moreover, it seems that when MPS is severe, a widespread pathology may manifest like a fasciomuscular armor under the skin (i.e., “fascial armoring”). For example, immunohistochemical examination of sampled fascia from low back pain patients demonstrates a myofibroblast density comparable to that found in “frozen shoulders” [14]. Activating many active loci simultaneously, by nature, might act to sympathetically “shield” an organism from acute threats at the expense of a fascial change. If tension is not released afterwards, fascia will be remodeled in that state and symptoms (e.g., muscular tension) will become chronic. Only, this muscular tension will not be muscular in its essence. This suggests that an acute sympathetic activity can have lasting effects on the organism’s fascia if not treated early after the acute event.

People with MPS fail to relax muscles due to a mechanical reason. Myofibroblasts can synthesize α-SMA fibers, which would transform fascia into a pseudo-muscular contractile tissue. It is not necessarily innervated and can remain contracted for years [51,83]. Since this tension is not innervated by voluntary control nor by motoneurons, and despite involvement of the SNS, seeing these patients as people who just “need to relax” as first line management is counterproductive. Failure to address the mechanical aspect of myofascial pain is expected to lead to a detrimental cycle of pain and over medicating (a cycle seen in studies addressing ineffective treatments) [11,84–86]. Also, if the pain in “non-specific low back pain” arises due to a fascial pathology, then calling it “non-specific” could be misleading.

Finally, sedentary behavior and active lifestyle are widely emphasized in public health. However, much focus is dedicated to cardiovascular and metabolic aspects. This review sheds light on the link between lifestyle and pain and suffering. It is worth noting that the most common recommended treatment of MPS found in this literature review is mechanical, including needle insertion, movement (passive or active, breathwork and vibration), and massage/manipulative therapy. It helps treat MPS or reduce fascial related pain [2,3,4,9,11,43,54–57].

#### Therapeutic considerations

Needling was found as a recurrent modality in literature. Needle treatment of TrPs increases pressure pain threshold [49]. Three elements might explain how needling can help treat MPS; These are (i) tensegrity mindset (ii) trigger points and satellites (iii) myofibroblast generated tension and stress shielding.

Since needle insertion itself seems to relieve pain [87], this may indicate needling creates a focus for a mechanical tearing action on the fascia. Meaning, the tension and shear stress inside fascia would pull on the weak point of fine needle insertion over time, until the sum force vectors, or horizontal components, at that point is eliminated or needle is removed. Cutting off tension and fascial fibers is expected to cause cells to rapidly contract, and subsequently lose their stress fibres and adhesion complexes [51]. If proto/myo/fibroblasts are present, as a node or a line (e.g., similar to the band described in pp. 23 Travel, Simons & Simons [9]), tearing fascia would affect the tensegrity structure. Mechanical stimuli can induce myofibroblasts stretch activated calcium channels, and calcium is coupled to contraction in these cells [60,61]. After calcium influx, MLCK activity might lead to increased contractile forces and tension acutely, if they do not lose their stress fibres and adhesion complexes. If needles tear the fibers, tension will decrease. Studies observe increase or decrease in contractility and stiffness, and witness “needle grasp” when inserted [88–91].

Myo/fibroblasts create tension in the attached matrix, and upon release of tension cells undergo contraction leading to a rapid contraction of the collagen matrix [51]. So, it may be that any “damage” by the needle actually allows for cutting off tensions between tense nodes in the network of (myofibroblasts) TrPs and satellites, mechanically inducing changes [88,91], and allowing for the returning of fascia to its place. If inserted deep enough, it might also free edematous fluid, HA, or other factors trapped within the layers (e.g., serotonin) [12,92,93]. This would gently change the dynamics of the structure and the entropy (hidden tension) of the system. Upon complete freeing of tension cells undergo apoptosis with reduction of α-SMA [94]. Studies indicate needling lowers myofascial stiffness as measured by shear wave elastography [95–97], suggesting a potential for a mechanical “re-alignment” of the fasciomusculoskeletal system and actual tensional release.

The more stress-shielding is present at the locus of insertion, the longer it should take for relief to happen. Stretch lesions along sensory nerves should be relieved too. This treatment acts like breaking the connections in a geodesic dome of the tensegrity model and then allowing for new healthier connections to form by natural remodeling. (The following video observations may help grasp this model [98–101]) Eliminating one node is not enough in this framework, on the contrary: if done improperly, it can only add tensions to other nodes in the structure and exacerbate the general pathology. Starting the relaxation process away from the focus of pain makes sense in this framework. It slowly and progressively begins to relax the primary, secondary, tertiary etc. nodes in the satellite network, approaching the highest area of unshielded tension later and collapsing the tension in a concentric way. According to the biotensegrity model, the peripheries carry tension affecting the focus and are equally important. Li et al. [102] state that currently available needle delivery systems deform and move soft tissue and organs. This may suggest that inserting a needle causes windup of connective tissue and modulation of the fascial network based on internal forces. Since fascia is made not only of proteins but is also composed of cells, the tensegrity dynamics will not be purely dictated by mechano-physics. “Tensegrity” is an oversimplification for this model. Alignment of the system should be done properly. The question is: What is properly? What is proper needling? Where, in which order, depth and time, is best for which symptom? Studies suggest needling affects facia at the point of insertion and at a distance [91,103–105]. How fast should the fascia be released? Indeed, it would be beneficial if we could monitor the mechanical state of fascia by palpation. By releasing the tension properly, the tissues can return to their healthy alignment without other areas exerting pulling forces. Following the “damage” of needling, the fascia will regenerate via myo/fibroblasts matching the bio-mechanical state. Evidence suggests that fascia regenerates within approximately 3-24 months after fasciectomy/fasciotomy [106,107]. It is unlikely that tearing the fascia (if done properly) will seriously compromise physiology as this would have manifested as a complication after every invasive surgical operation. New extracellular fiber deposition will be based on optimized structural conformation [22]. Some may say inserting several needles subcutaneously sounds like a surgical intervention of the fascia. A gross illustration of this framework can be found in S1 Fig 1A-D in the supplementary material (S1 Fig 1A-D).

#### Demonstrating the model of “Fascial Armoring” with empirical findings

Examples in support of the suggested mechanism may have transpired in several studies. Three findings are briefly presented as follows. A study of 37 patients who were all diagnosed with plantar fasciitis and all had been treated with corticosteroid injection into the calcaneal origin of the fascia [108]. All had a presumptive diagnosis of plantar fascia rupture. 30% described a sudden tearing in the heel, while the rest had a gradual change in symptoms. Most of the patients had relief of the original heel pain, but it had been replaced by a variety of new problems, including dorsal and lateral midfoot pain, swelling, foot weakness, metatarsal pain, and even metatarsal fracture [108]. Symptoms tended to localize to the dorsal side of the foot [108]. In all 37 patients there was a palpable diminution and footprints showing flattening of involved arch, and MRI showed fascial attenuation [108]. The study’s author concluded that plantar fascia rupture had occurred. The majority had resolution of the new symptoms within 12 months. In the rest, symptoms remained [108].

A second study encountered the same enigma: 68% of patients with plantar fasciitis had a sudden rupture associated with a corticosteroid injection. Although it relieved the original pain, these patients developed new problems: arch or midfoot strain, lateral plantar nerve dysfunction, hammertoe deformity, and stress fracture [47].

These examples might suggest there is room for a more sophisticated strategy in treating the fascia instead of simply aiming and firing at the pain. Maybe this should be interpreted not as an adverse event, but as a testimony that the treatment modality undermines the tensegrity system. Conceivably, behind this is a release of fascia in an area of very high tension, causing too drastic of a change in the tensegrity structure, instead of approaching this tension gradually. A sudden change might shift the forces to other areas and exacerbate the imbalances [109], perhaps sufficient to cause bone fracture. The mechanism by which the complications resolved over a period of 12 months is expected to happen via ECM remodeling and stress shielding the foot from tensegrity tension. Moreover, it was found that steroid injection decreases fibromatosis, fibrosis, and myofibroblasts in adhesive capsulitis [110], suggesting injection to the plantar fascia has potential to affect myofibroblast generated tensegrity forces.

The third finding that might be interpreted under these principles, is the phenomena of: “following a tooth extraction, a pain behind the ear and on the side of the face in the day or so prior to facial weakness often constitute the earliest symptom of Bell’s palsy” (Kasper et al. 2018, p222-223) [3]. Guided with a tensegrity mindset, this may suggest a relationship in the “geodesic” face: a pain develops behind the ear possibly as a “reciprocal” point for tension in the jaw, which happens to be near the emergence of the facial and trigeminal nerves from the skull. This is a reasonably susceptible area with curved bones to serve as a focus for tension. Of course, it does not necessarily mean that releasing tension from around the mastoid process is the key to preventing the complication. Tensegrity domes are made of many nodes, and “to change a node is to change the dome”. Therefore, pain behind the ear is first and foremost an indication of extreme tension, and only then pain. A tension like every other myofascial tension, e.g., plantar fasciitis. However, this pain is located at a dermatome of a cervical root, which is a distant nerve and is unlikely to be directly injured by tooth extraction. A sudden intervention in the face probably emulates the same action as a calcaneal injection. If high tension is present (for example a temporomandibular tension seen in up to 25% of the population [2]) it will have the same potential energy to damage structures in the face, as implied by a metatarsal fracture. In the face it seems to cause a palsy of a nerve, like the foot in plantar fasciitis, as discussed above. The degree of damage is modulated by the degree of the hidden energy [109]. If correct, this means two possibilities present themselves: either this palsy is not a Bell’s palsy, or a Bell’s palsy is not idiopathic. In any case, it highlights the instrumental importance of gentle and progressive release of fascia in a proper way. Therefore, “not feeling much” may be a good thing when needling fascia. In fact, we can predict sometimes pain might worsen at first, reflecting a non-ideal modulation of force vectors. Less pain during treatment can also lead to more compliance from patients. Research has yet to establish empirically if fascial release prior to (or even after) tooth extraction, can affect the rates of this palsy. Modulation of tensegrity vectors may be a factor in reported cases where (i) treatment of Dupuytren’s disease leads to Boutonniere deformity [111], (ii) carpal tunnel release increases risk of trigger finger [112,113], (iii) treating the elbow for tennis elbow requires a supposedly unrelated shoulder arthroscopic decompression [114], (iv) simultaneous bilateral digital flexor rupture occur [115], or (v) spine surgery leads to compartment syndrome of the foot [116]. It seems plausible since fascia is a system. Fascia appears to connect the distant hip and ankle joints, not only anatomically, but also mechanically, supporting the concept of myofascial connectivity [18]. Most skeletal muscles of the human body are directly linked by connective tissue [40]. Acute bout of stretching of the lower limbs increases the maximal range of motion of the distant upper limbs and vice versa [18]. Fascia is continuous from the trunk across the upper and lower limbs [18], and because of the direct morphologic relation of the hamstrings and low back region, relieving tension of the posterior thigh muscles could be a conceivable approach to alleviate back pain [117].

#### Epilogue to part 1

In conclusion, under the framework of tensegrity, it might be logical to first needle the dorsal foot and foreleg in plantar fasciitis, for example, assuming connective tissue is pulling the plantar fascia (i.e., weak link in the tensegrity structure) and causing pain. Therefore, aiming to primarily treat the underlying tensional abnormality of fascia, and through this improving the symptoms of pain. Actually, it seems a TrP with severe localized pain may not be the source of pathological tension because myofibroblasts stress shield their area from tension: fascia is a living tissue that pulls rather than pushes. Treating only the painful point is expected to lead to an endless chase of migrating pain whose location is determined by dynamics of the tensegrity structure. Tension is likely to focus more near hard and angled surfaces that have sharp force-gradients; initially generating active TrPs (if a nerve is there to experience this) that have yet to sufficiently stress-shield themselves with a fiber web. By this rationale we may expect points to be concentrated more near areas of fascia adjacent to solid or bony edges (spine, face, pelvis, chest, shoulders, knees, periosteum, perineurium, etc.). Tension will be present [118,119], but pain depends on the degree of sensory nerve involvement [120,121]. This understanding may put MPS and fibromyalgia on the spectrum of a common mechanism, whereby a more widespread pain and tenderness appears, at least in part, perhaps due to “archipelagos” of stress-shielded points and taut-bands that generate and mask extreme tension. If true, a severe manifestation of this might be described by some as equivalent to a sort of mild to moderate chronic compartment-like syndrome of the whole body. Which may explain why “MPS” can “develop” into “fibromyalgia”, at least in a subset of patients. A biological representation for this might be: myofibroblast begin to affect the body in a generalized MPS cascade and therefore transform the fascia into a fibrotic contractile tissue (perhaps with a reciprocal central sensitization). Recalling that dysfunction of the posterior layer of thoracolumbar fascia has been reported as a chronic compartment syndrome of the paraspinal muscles [26].

These conclusions may be part of the explanation to why these “functional” pain syndromes are epidemiologically so closely associated with one another (as observed in population studies [122,123]), why significant overlap is seen between various associated pain syndromes (reflecting anatomical overlap of involved fascia), why fibromyalgia tender points seem to distribute more near bone protrusions, why a study finds fibromyalgia symptoms and neuropathic pain correlate with tender point count (Amris et al. [121]), why chronic fatigue is a common manifestation, why routine inflammatory markers can be mostly silent, why imaging modalities fail to illustrate a clear familiar abnormality, why mechanically puncturing their fascia recurs in literature as an empirical treatment option for them, and why fibromyalgia manifests with extremely high intramuscular pressures and can be accidentally resolved by invasive surgery (as will be discussed below). It can also help explain histological and clinical evidence indicating a picture of widespread non-severe chronic ischemia. Studies of fibromyalgia observe various organic abnormalities [124–141] which might support this framework. To the best of the author’s knowledge, no study is found to describe the manifestation of MPS when global and widespread.

### Part 2 – Considering “fibromyalgia” as an entity involving mechanical compression

Part 1 of the discussion focused on the theoretical model of “fascial armoring” formed based on the empirical evidence found in the systematic search and scoping review. A myofibroblast network generating tensegrity tension (i.e., “fascial armoring”), when widespread, predicts further abnormalities. In this section empirical evidence is presented in support of fibromyalgia as a global chronic compartment-like syndrome driven by myofibroblast-generated-tensegrity-tension. Findings from various fields including cardiovascular, metabolic, musculoskeletal, integumentary, and surgical, are discussed.

#### Cardiovascular findings

Studies show tissue stiffness is significantly increased around TrPs [142], and blood vessels near TrPs have retrograde flow in diastole, indicating a highly resistive vascular bed [138]. A global compression might explain a few findings that will be mentioned here briefly. Various abnormalities in fibromyalgia suggest an underlying widespread organic pathology that impairs perfusion throughout the vascular system. Capillary microscopy studies of fibromyalgia show fewer capillaries in the nail fold and significantly more capillary dilatations and irregular formations than the healthy controls [128]. The peripheral blood flow in fibromyalgia patients was much less than in healthy controls [128]. This suggests that functional disturbances are present in fibromyalgia patients [128]. While one study shows a decrease in transcapillary permeability in fibromyalgia patients [129], few studies find abnormalities in the arterial system as well [143,144]. However, studies of peripheral arteries in fibromyalgia are scarce. Findings from a study of biopsies of fibromyalgia cases without obvious muscle trauma indicate definite but nonspecific muscle changes which are suspected to be secondary to chronic muscle spasm and ischemia of unknown etiology [124].

Katz et al. [145] found intramuscular pressure measures substantially higher in the trapezius of patients with fibromyalgia with a mean value of 33.48 ± 5.90 mmHg. Only 2 of 108 patients had muscle pressure of less than 23 mmHg. The mean pressure in rheumatic disease controls was 12.23 ± 3.75 mmHg. The burden of the pressure abnormality may help explain the diffuse muscle pain of fibromyalgia and may be an intrinsic feature of fibromyalgia [145]. Therefore, fibromyalgia as a disorder of exclusively central pain processing should be revisited [145]. Substantially higher muscle pressure aligns with an analogy of “a sort of mild to moderate global chronic compartment-like syndrome” which either compresses the muscles or distends the muscle spindle, or both, and may explain why anxiolytics and muscle relaxants do not work very effectively with this entity. At the end of arterioles, mean pressure is approximately 30 mmHg, and then decreases further in capillaries and venules [146]. Chronic intramuscular pressure of 33.48 ± 5.90 mmHg might reflect some abnormality that ultimately affects starling forces significantly, as well as the pressure gradient along arterioles, capillaries, and venules. The most widely used diagnostic criteria for diagnosing chronic exertional compartment syndrome (CECS) are those proposed by Pedowitz et al. for CECS of the legs [2] and include at least one of the following intramuscular compartment pressure measurements: (i) pre-exercise pressure ≥15 mmHg (ii) 1-minute post-exercise pressure of ≥30 mmHg, or (iii) 5-minute post-exercise pressure ≥20 mmHg. Therapeutically, reduction of muscle pressure may change the clinical picture substantially in fibromyalgia [145]. If myofibroblasts are indeed the generator of tension, then abnormal muscle pressure should be reduced by releasing the fascia, according to the “fascial armoring” model, since it is currently not a psychologically generated contraction. If fascial compression impedes starling forces and causes low-grade ischemia, it might be biologically expected that the body will secrete factors that increase total number of capillaries and venules, and factors that vasodilate veins (e.g., nitric oxide). Such a biological response would lower total venule resistance via parallel resistance and re-establish a pressure gradient in the vessels.

#### Metabolism related findings

A microdialysis study examined interstitial concentrations of metabolic substances in fibromyalgia [147]. Concentrations of lactate, glutamate, and pyruvate were significantly higher in patients compared to controls [147]. A study by Mclver et al. [148] found that during resting conditions, the ethanol outflow/inflow ratio (inversely related to blood flow) increases in fibromyalgia patients over time compared to healthy controls (p-value <0.05). Fibromyalgia also exhibited a reduced nutritive blood flow response to aerobic exercise (p-value <0.05). There was an increase in dialysate lactate in response to acetylcholine in fibromyalgia [148]. A recent genomic study found the presence of a C allele at the single nucleotide polymorphism of mitochondria m.2352T>C was significantly associated with increased risk for fibromyalgia (OR=4.6) [149]. It was linked to decreased mitochondrial membrane potential under conditions where oxidative phosphorylation is required. The study suggested that cellular energy metabolism contributes to fibromyalgia and possibly other chronic pain conditions, indicating a role of oxidative phosphorylation in pathophysiology of chronic pain [149]. A clinical trial found hyperbaric oxygen therapy can lead to significant amelioration of all fibromyalgia symptoms, with significant improvement in quality of life [150]. Fibromyalgia has a hypomethylation DNA pattern, which is enriched in genes implicated in stress response and DNA repair/free radical clearance [151]. From the discussion above, the link between fibromyalgia and oxidative stress, increased intramuscular pressure (that is higher than all three criteria for CECS by Pedowitz et al.), and reduced blood flow measurements, is self-explanatory.

#### Muscular stiffness and the spine

Wachter et al. [131] quantified muscle damping, which reflects muscle tension, in fibromyalgia patients, and found all patients had increased muscle damping, some in both legs [131]. Mean values were more than twice that of controls and maximum values more than three-fold higher [131]. The increased DNA fragmentation and ultrastructural changes in muscles of fibromyalgia patients suggests it may be a result of chronic contraction [125]. Wachter et al. stated increased muscle tension is not mediated by extrafusal muscle fibers and that the alpha and gamma motoneurons are not involved in those contractions [131]. They suggest stiffness or tension may be due to sympathetic activity, however, they state this explanation does not account for why increased damping is seen more often in one leg rather than in both [131]. TrPs appear on ultrasound as focal hypoechoic regions and with reduced vibration amplitude on vibration sono-elastography, indicating increased tissue stiffness [138]. A non-symmetrical tensegrity abnormality could account for why tension can be mediated not by alpha or gamma motoneurons or extrafusal fibers, and at the same time unilateral.

Other body structures are also involved, among them are nerves. Perez-Ruiz et al. [152] found high rates of carpal tunnel syndrome in fibromyalgia. Why is a nerve compression syndrome found to be more common in fibromyalgia, if it does not reflect some compressive abnormality? (Fascio)musculoskeletal symptoms are reported often by patients: stiffness is one of the most common complaints [153,154]. A survey of 2,596 fibromyalgia patients finds patients ranked the intensity of their pain lower than the intensity of morning stiffness [153]. Considering central sensitization as a sole mechanism raises several questions: why is morning stiffness so commonly reported among fibromyalgia patients, and even ranked higher than pain? Is it simply because of sensitivity, stress, and physical deconditioning? Does central sensitization occur only in certain sensory tracts like pressure, proprioception, and pain? There seem to be little to no reports of extreme sensations of vibration, for example. Are the locations of central sensitization random, or is there some sort of pattern? Painful points seem to be distributed in the same topography among different individuals. However, if the localization of pain and hyperalgesia is not random, why can sensitivity appear in almost any area in the body? Why are these areas not correlated with the homunculus? How does pain migrate? And what is special about the occiput, second costochondral junctions, knee pad, and two centimeters distal to the lateral epicondyle - points used to diagnose fibromyalgia (as described by Kasper et al. 2018; pp. 2637 [3])?

Sugawara et al. [155] suggest mechanical compression of the dorsal root ganglion (DRG) by a mechanical stimulus lowers the threshold needed to evoke a response and causes action potentials to be fired. Action potentials that may even persist after the removal of the stimulus and high mechanical sensitivity are suggested by this (in vitro) study. More in vivo and in vitro studies suggest the same [156–158]. It was noted that dysfunction of the thoracolumbar fascia has been described as a chronic compartment syndrome of the paraspinal muscles [26]. It is also plausible that pulling would also have a similar effect on the DRG.

Wang et al. [159] find a bidirectional association between fibromyalgia and gastroesophageal reflux disease (GERD). Trying to explain this without a mechanical peripheral element seems difficult. How do current theories explain this bidirectional linkage? It is possible GERD carries a strong overlooked psychological component, but maybe we should give fibromyalgia more mechanical credit. This entity, if it is what it seems, would probably affect gastroesophageal pressures and relaxations, e.g., DRG and the sympathetic chain against a vertebral body. If compression has a similar effect on the sympathetic ganglion/trunk as it does on the DRG, it might add a component of sympathetic abnormalities. Some studies suggest mechanical pressure on the sympathetic trunk due to osteophytes can cause various abnormalities [160]. Studies suggest para-articular fibrosis in the spine can involve neural structures and cause tethering and irritation of nerves [161]. A compressive factor can explain decreased pressure/pain thresholds and explain why they happen to be strongly associated with autonomic abnormalities. It also implies these abnormalities can be illusive and reversible in certain conditions, as will be discussed below. By the same rationale we might expect higher incidence of obstructive rather than central sleep apnea, even after correcting for body mass index.

#### Complete resolution of fibromyalgia

Several studies point to a peculiar phenomenon: Saber et al. [162] witness high rates of complete resolution of fibromyalgia following laparoscopic Roux-en-Y surgery. By which mechanism does a relentless longstanding central sensitization completely resolve following abdominal surgery? Could this be a result of an overlooked fasciotomy? The authors of that study suggest resolution may be due to weight loss, increased physical activity, and lifestyle changes. However, this idea does not align with findings of Adkisson et al. [163]. They studied the effect of parathyroidectomy on fibromyalgia among 76 patients diagnosed by specialist or primary care physician. Findings show that 21 percent of fibromyalgia patients discontinue all fibromyalgia drug medications after parathyroidectomy, and 89 percent have relief of one or more fibromyalgia symptoms postoperatively. These changes occurring as early as one week after neck surgery [163]. Yet, 11% of patients had no improvement in symptoms at all. The authors explain the complete resolution in 21 percent by suggesting a misdiagnosis, i.e., these patients actually did not have fibromyalgia in the first place. However, if this was true, we might expect these individuals to cluster more towards the group not diagnosed by a specialist. Surprisingly, 23 out of 76 were diagnosed by a rheumatologist, out of them 96% had symptom improvement, and 70% were able to discontinue all medications after parathyroidectomy [163]. Uncannily, 88% of those who did not experience any improvement in fibromyalgia symptoms were diagnosed by a primary care physician (p-value <0.05) [163]. The statistics suggest some non-random phenomenon caused a bimodal-like distribution. In light of this study, it might be hypothesized fibromyalgia is related to a parathyroid cause. However, a similar enigma is found with irritable bowel syndrome (IBS), a functional disorder with no definite organic findings and closely associated (or overlaps) with fibromyalgia. Some suggest fibromyalgia and IBS are actually the same entity [154]. IBS is shown to be relieved below Rome II criteria in 80 percent of patients after laparoscopic fundoplication for GERD [164]. Looking for a common denominator one can hypothesize that relief is achieved due to intense pain exposure and general anesthesia that resets the brain circuits, somewhat similar to electroconvulsive therapy combining gate control. But it does not align with findings that tonsillectomy increases risk of IBS [165], that umbilical hernia repair surgery seems to predispose to IBS [166], and that patients with fibromyalgia have a higher incidence of suffering IBS after appendectomies [167]. Laparoscopic cholecystectomy has been shown to deeply influence fibromyalgia symptoms [132]. Moreover, hysterectomy, with or without oophorectomy, seems to worsen fibromyalgia [168].

Oddly, why does the location and type of surgery seem to regulate the positive or negative impact on these somatic conditions? Indeed, it was suggested in this literature review that MPS/TrPs can cause chronic changes in bowel habits. Should we suspect that in these associated disorders, the connecting motif is connective tissue? This may imply that the location of the surgery has different effects on fascia, even remotely, due to- and regulated by-tensegrity forces. The lateral raphe and common tendon of the transversus abdominis are suggested to form connections between, and transfer tensions between the abdomen and the paraspinal muscles and sheath [14,169]. This can explain how laparoscopy might release a compression on the DRG. Interestingly, this would mean the clustering of patients experiencing benefit post-parathyroidectomy towards the group diagnosed by a rheumatologist, and those not experiencing benefit towards the group diagnosed by primary care physician, isn’t due to misdiagnosis but precisely because rheumatologist really identify those with fibromyalgia statistically significantly more accurately.

Now, what is missing from current theories about fibromyalgia? On one hand, 21st century’s best physicians and psychotherapists are not enough for fibromyalgia to relinquish, while on the other hand, parathyroidectomy is. What do the surgeons do, even without special intent, that we haven’t tried? How to see that which is being overlooked? It appears this entity, or the tissue that is the veil, is responsive to surgery more than to opioids and anti-epileptics.

According to central sensitization, is a parathyroidectomy able to fix the pain matrix? And can hysterectomy impair it? What about below knee amputation?

#### Multiple theories for one medical puzzle?

More studies indicate fibromyalgia may include also easy bruising [154,170], hair-loss [154], reduced skin innervation [171], tingling, creeping or crawling sensations [172], reduced fecundity or infertility [173,174], urinary urgency [164], changes in bowel habits [3], decreased optic disk perfusion [127], dry eyes [172], dry mouth [154], hearing difficulties [154], functional voice disorders (including muscle tension dysphonia) [175], wheezing [154], seizures [154], impaired cognition [154], and Raynaud’s phenomenon [154,176].

There are several theories trying to explain fibromyalgia including: central sensitization and neuroplastic pain circuits, autonomic dysfunction, neuroendocrine and stress, social and psychological (including personality and cognition), and several more theories. Can they be the explanation of all these manifestations? Do psychosocial theories or somatization explain these? No single theory seems to fully account for fibromyalgia. For example (some of the questions below were already answered, others will be answered later while others remain to be answered by further research):

##### If it is sympathetic-parasympathetic dysregulation

why is muscle damping found to be increased unilaterally more often than bilaterally, why is postural hypotension not in the diagnostic criteria, and why is it strongly associated with sicca but lacks reports of chronic sialorrhea? Why urinary urgency but not retention, and how to explain empirical findings of autoantibodies?

##### If it is inflammatory or autoimmune

where are the standard inflammatory markers, why is it so closely associated with irritable bowel syndrome of all conditions, and why does it respond to tricyclic antidepressants? What do the occiput, scapula, and knee possess, for example, that is absent from all interphalangeal joints and parietal bones? Are bone ridges a necessary-and-not-sufficient condition for tender points? What underlies the close association with acquired immunodeficiency syndrome [177], and why does a study find placebo has a better outcome than steroids as a treatment for fibromyalgia [178]? The reason pharmacological treatments require prudence is because systemic drugs would distribute in an imbalanced tensegrity structure equally throughout, which can be an inherent disadvantage in this case. According to this framework, systemic steroids are expected to indiscriminately modulate the tensegrity structure, supposedly arbitrarily leading to an exacerbation or relief, depending on the structure. More specifically, the success of some pharmacological agents that target myofibroblasts will depend on the balance between pro-survival signals (e.g., mechano-transducing signals) and pro-apoptotic signals.

A low-grade neuroinflammation might occur sub-clinically in fibromyalgia, but what drives it? The full cytokine profile of MPS and fibroblasts is not covered in this review, however, some studies suggest chronic myofascial pain is associated with, or causes, elevation of cytokines and inflammatory mediators, fibroblast growth factors, serum reactive oxygen species, and neuroendocrine signaling [19,179–181]. However, not all studies find the same [181]. Fibroblast from fibromyalgia patients show a differential expression in proteins involved in the turnover of the ECM and oxidative metabolism that could explain the inflammatory status of these patients [182]. Biopsy studies of fibromyalgia patients find that, in peripheral fibroblasts, TGF-β gene expression is significantly higher in fibromyalgia patients compared to controls [130]. If one accepts only alternative theories: why and how is TGF-β expression two to three-fold higher in peripheral fibroblasts of fibromyalgia patients (p-value <0.001)?

Is there any evolutionary sense for support of fibromyalgia as autoimmune? Cancer is rigid and is affected by connective tissue rigidity [16]. Suppose tissue stiffness regulated antigen presentation, somehow, as a way of the body to defend itself from pathological rigidity, what would be the implications for fibromyalgia based on tensegrity dynamics? If this is part of the explanation for autoantibodies in fibromyalgia, there is at least some evolutionary background for the autoimmunity part of it. Substrate stiffness influences phenotype and function of human antigen presenting dendritic cells and affects B-cells, macrophages, and other immune cells [183]. Myofibroblast can engulf their surroundings [184], and in certain conditions are able to present antigens via MHC class II to T lymphocytes [185–188]. A study from 1990 of 20 patients with fibromyalgia suggests the majority have anti smooth muscle antibodies [189]. Such a defensive mechanism would be successful in selection throughout generations only if it allowed for some autoimmunity, in order to target the suspicious cells. Compromising self-recognition was probably beneficial rituximab does not do the whole job, maybe some clone population keeps presenting antigens.

##### Central sensitization is the most accepted explanation

however, there are several questions that this theory does not seem to fully explain and some empirical findings that the theory did not seem to predict. How is fibromyalgia mechanistically different from “tension-type-headache”, “nonspecific-low-back-pain”, “nonspecific-neck-pain”, and “MPS”? What is the pathophysiological difference between “primary fibromyalgia”, “secondary fibromyalgia”, “secondary-concomitant fibromyalgia”, and “juvenile fibromyalgia”? Is fibromyalgia the same entity or different from “chronic widespread pain”? In terms of “fascial armoring”, there is no need to multiply entities, it remains myofibroblast-generated-tensegrity-tension, though, each tensegrity is different, anatomically. The fascial armoring model, intrinsically, has variations. Why does “chronic fatigue syndrome” (CFS) manifest with pain in tender points? Since fatigue is the predominant symptom rather than pain, it might indicate that deeper layers of connective tissue are more involved. How does the pathophysiology of fibromyalgia overlap with that of headaches and CFS, if searching for it only within the cranium? The boundary of an entity goes as far as the boundary of the cellular process that is behind it.

Can central sensitization be resolved? If so, what is the modality and mechanism of action of the cure? In the long term, do opioids worsen the pathology? According to central sensitization, why prefer milnacipran, duloxetine, and pregabalin, but not venlafaxine, tramadol, or gabapentin? Is diet important? A pharmacological or nutritional agent that induces myofibroblasts is expected to be either ineffective or harmful in the long term in fibromyalgia. From the standpoint of “fascial armoring”, an antidepressant that induces myofibroblasts might have some positive side effects, e.g., improved mood.

Will surgery exacerbate central sensitization, relieve it, or neither? What are tender spots and how does the topography remain conserved between individuals? These spots seem fundamental because fibromyalgia can be diagnosed based on them. Why two centimeters distal to the lateral epicondyle and not midway between the elbow and wrist? Will this topography change if there are congenital deformities in the brain/spine? What if a deformity in the elbow? Which is the constant that allows for the preservation of this point distribution? How do we know there is no peripheral organic cause for the pain if we haven’t looked at tensegrity? Is “post COVID-19 syndrome” similar, different, or is it the same? If the same, according to this model, COVID-19 should induce myofibroblasts. Which comorbidities interact best, and which are the worst for fibromyalgia, based on central sensitization, theoretically? One might offer multiple sclerosis, tetraplegia, adrenomyeloneuropathy, Cushing’s disease, or congenital adrenal hyperplasia. So then, why is Ehlers Danlos/hypermobility syndrome so closely related to fibromyalgia?

The pathogenesis of peripheral and/or central nervous system changes in chronic widespread pain (CWP) is unclear, though, peripheral soft tissue changes are implicated [190]. Some evidence from interventions that attenuate tonic peripheral nociceptive impulses in patients with CWP syndromes like fibromyalgia suggest that overall fibromyalgia pain is dependent on peripheral input [190]. Allodynia and hyperalgesia can be improved or abolished by removal of peripheral impulse input [190]. Central disinhibition is also hypothesized. However, this mechanism also depends on tonic impulse input, even if only inadequately inhibited [190]. Thus, a promising approach to understanding CWP is to determine whether abnormal activity of receptors in deep tissues is part of the manifestation and maintenance of this condition [190].

Perhaps the relationship between fibromyalgia and hypermobility syndrome has to do with changes in ECM properties that induce myofibroblasts, thus initiating a widespread MPS cascade? Studies of patients with Ehler-Danlos/hypermobility syndrome find up to 70-100% have CWP and evidence supports a close association between hypermobility syndrome and fibromyalgia [191,192]. Of all associations with fibromyalgia, Ehlers-Danlos/hypermobility syndrome is one of the closest, perhaps even closer than depression. Why? Studies of hypermobility syndrome find cells express an increased transition to the myofibroblast phenotype [193,194]. A study suggests changes of collagen microarchitecture regulate myofibroblast differentiation and fibrosis independent of collagen quantity and bulk stiffness by locally modulating cellular mechanosignaling [195]. The indistinguishable phenotype of myofibroblasts identified in hypermobility spectrum disorders resembles an inflammatory-like condition, which correlates well with the systemic phenotype of patients [193]. These findings suggest that these multisystemic disorders might be part of a phenotypic continuum rather than representing distinct clinical entities [193]. According to this discussion, widespread induction of myofibroblasts suggests a possibility for generalized peripheral pain caused by generalized tensegrity tension of collagen/elastane is somewhat anticipated based on fascial biology. Even at the usual shear modulus of fascia, in this population joint laxity might allow tethered nerves to be affected more than normal. The above suggests that finding only a “normal” range of motion in the joints of a hypermobile individual is abnormal and pathological not only because it restricts their movement, but because it is a marker for a diseased fascia.

##### Psychological and social theories

evidence for the input of psychological/psychosocial factors and stress in the pathogenesis of fibromyalgia is suggested in studies [154]. Cognitive behavioral therapy is recommended in cancer pain, but we obviously do not conclude that cancers are psychosomatic. Adverse childhood events are associated with increased risk for diabetes mellitus type 2 [196], yet only a rare few define diabetes mellitus as psychosomatic. So, if fibromyalgia is caused by stress and low mood or due to childhood experiences of distress, how does it reduce skin innervation and optic disc perfusion, and why selective serotonin reuptake inhibitors (SSRIs) or anxiolytics are not the first line treatment? If traumatic memories can maintain the pain of fibromyalgia, do strokes or Alzheimer’s disease relieve it? If abnormalities in cortisol levels can be enough to lead to such a clinical picture, why aren’t they enough to cause even mild hyperglycemia, neutrophilia, or thrombophilia? No osteoporosis? Hyperpigmentation? And no salt craving or hyponatremia. If cortisol has a major role, why is Cushing syndrome not usually portrayed with morning stiffness and pain in 18 arbitrary points (arbitrary from a neurological point of view) that are all adjacent to bone protrusions and are also adjacent to strong muscles and fascia pectoralis, muscles of mastication etc.)? Is there any outstanding “fibromyalgia-ness” in either Adisson’s or Cushing’s disease, or in Pheochromocytoma? Endocrine entities involving these hormones are not entirely free of emotional distress either.

Psycho-emotional explanations are a double bind for fibromyalgia: if muscle tensions and contractions are psychologically induced by mental stress, how are they maintained chronically with such high intramuscular pressures if patients are often fatigued? On the other hand, if habitual psychological muscle contraction is what leads to the fatigue, why can’t they relax, and how do muscles contract independent of alpha and gamma motoneurons and extrafusal fibers? Would we not expect a combination of muscle relaxants and meditation to be more efficacious? What kind of an entity can be both systemic and unilateral? A sympathetic theory does not suffice, as stated by Wachter et al. [131]. S1 Fig 1E illustrates a possible solution for a systemic yet seemingly “unilateral” pathology.

Is depression a causal factor, and if so, why are antidepressants not more effective? Is there a global abnormality in tryptophan’s derivatives? Does physio-emotional Stress cause fibromyalgia, or does fibromyalgia cause physio-emotional Stress? At which stage, approximately, does the mechanism of this entity diverge from that of Stress, and why is it closely associated with Sjögren’s syndrome? Why pass over pancreatitis? And is there a prediction for the interaction with asthma, for example? The author could not think of an evolutionary rationale in having a self-destructive infinite positive-feedback of pain in two to six percent of a species, with no peripheral organic lesion, due to history of emotional trauma, or due to any reason. These entities were designed throughout evolution, no? Life must go on and we need to move for life to move on. Cancer uses fetal genes to multiply, cystic fibrosis is part of variety and selection, allergies are immune cells being overprotective, iron deficiency can cause pica, and COVID-19 is a big accomplishment for viruses. Why would the brain amplify pain with no organic lesion? Evolution might be offended; and treating that malady with exercise is not all intuitive either. The human mind is most complex but is not most frail. Is connective tissue a non-issue? Why is the focus specifically on pressure and pain? Does psychology navigate sensitization towards predetermined sensory tracts, or is there genuine sensory input of pressure arising from the periphery and causing central neurological adaptations? If the former-why those tracts? And if the latter-what drives the force that causes pressure in the periphery? The character structure? The tensegrity structure? Myofascial tissue? Does the reader know of any entity that can sustain generalized mechanical pressure with no obvious cause? For the sake of a scientific discussion, let us include any hypothetical entity as well.

If fibromyalgia carries such a strong psychological component, why did a study find an odds ratio of 2.53 and effect size of only 0.51 for anxiety disorder in fibromyalgia, while for fasciitis and muscle spasms it found odds ratios of 5.27 and 6.05, with an effect size of 0.92 and 0.99 (p-value <0.0001), respectively [197]? Does fibromyalgia as a “functional somatic syndrome” deserve some re-evaluation, and are we to dismiss these statistics as not being empirical clues? Why did a study find fasciitis to be one of the three most associated conditions with fibromyalgia, while anxiety was not? Is there, or is there not, a problem with the fascia of fibromyalgia patients? An odds ratio and effect size approximately double that of anxiety disorder, but if the fibromyalgia? How to interpret the topography, then? Is there anything we can learn from the pattern? Are the points entirely psychological, are they created in the central nervous system, or does the theory require a peripheral mechanism to fill this gap? Which is largest – the variance of fibromyalgia’s prevalence between different societies, between the big five personality traits, or the variance of prevalence between collagen polymorphisms? Does “post-traumatic anxiety syndrome” predispose to fibromyalgia, or are they overlapping conditions? What sets the boundary of this entity? ‘Psychology’ can be infinite. Why not pica too? Is there any determinism? Does fibromyalgia originate from the head or from the body? And if it comes down to the mind-how does the collagen polymorphism make one so much more sensitive to psychological problems? What can we learn about a fascial pathology if it affects the mind, in theory? By which mechanism does a psycho-neuroplastic pain cause so much muscle spasms but no abnormal Babinski sign, no clasp knife spasticity, no paralysis, no fasciculations, no hyperreflexia, no sensory loss, no ataxia, no dystonia, no atrophy, and no clonus, and why are these spasms different? Even if this shadow hides in the subconscious or elsewhere it needs to have some way to get to the muscle. Are these spasms driven by a psycho-neurological pathology of the central nervous system that contracts muscles without using extrafusal fibers or motoneurons, or that activates a unilateral sympathetic response, or should we explore unexplored rheumatological avenues?

##### “Somatic” needs clarification

Is “fascial armoring” part of “Somatic”? There must be some pathophysiology to it because it obviously is not magic and any model that tries to tackle fibromyalgia’s pain cannot be shy of “Somatic”, too. To rely on ”using problem, no? Having schizophrenia seems to be different and unfortunately searching only inside cranium limits our options. (The term “inside cranium” is used because the brain is certainly not alone in it, the meninges is there, too.) When excluding magic, imagination, and schizophrenia, what are we left with? A ghost in brain, or a shadow in fascia? Science is free to choose where best to look. Gupta et al. [198] studied CWP in somatising individuals. They conclude that while a high tender point count is associated with the onset of new CWP, a low pain threshold at baseline is not [198]. With our new view of MPS it is logical, since tender point count would be a marker for a fascial tensional pathology, leading to widespread pain and a lower pain threshold later.

Why and how does MPS develop into fibromyalgia? Is MPS also psychosomatic? Essentially, any answer to that would depend on our definition of “psychosomatic”. If “psychosomatic” has a peripheral factor involving myofibroblasts, then, the answer is: probably yes. Therefore, this model suggests that between a treatment that focuses mostly on the brain or one that focuses mostly on the tensegrity, the one focusing also on the tensegrity should be more effective, at least in the medium to long term. In other words, a combination of cognitive behavioral therapy and exercise with any current pharmacological treatment protocol, should be less effective than, for example, a combination of movement therapy, hyperbaric oxygen chamber therapy, psychotherapy, and minimally invasive therapy (tensegrity based needling). It also suggests that concentration of myofibroblasts and α-SMA in involved connective tissue should correlate better with the Fibromyalgia Impact Questionnaires than do frequency of pain catastrophizing or severity of emotional trauma. Has the reader ever felt an itching sensation, headache, or repeated muscle twitching, with no clear reason? Was it ever psychosomatic?

There are those who suggest fibromyalgia is not a real entity or that it is caused by sleep disturbances [154]. However, the relatively modest efficacy of antipsychotics and/or melatonin and/or z-drugs implies it is probably neither an imaginary hallucination nor due to lack of sleep. If it truly is only pain catastrophizing due to low psychological resilience, then animal models are probably futile, and why are women more affected than men?

Noteworthy, two parallel mechanisms (or more) might be at work and are not necessarily incompatible with each other. Notwithstanding, if the mechanism starts in the fascia, and if we rarely study the fascia in fibromyalgia, how will we find the mechanism?

How is it that multiple theories, even when combined, do not seem to fully explain this entity? Is there any tissue out there that all theories have overlooked? A tensegrity model is not without its questions: (i) How are depression and Lyme disease linked, and why is there wheezing, insomnia, and small fiber neuropathy? (ii) Self-mutilation? [199] (iii) Do antidepressants affect myofibroblasts? (iv) How is the brain involved? (v) Did biopsy studies in fibromyalgia find abnormalities in peripheral myo/fibroblasts, α-SMA, or TGF-β? (vi) What is altered if myofibroblasts have chemotaxis? (vii) If the tensegrity’s state can be different in each individual, how to minimize false negative results in research and how to tailor the treatment accordingly? (viii) What if the “armor” develops in the deep fascia more than the superficial? (ix) What if an imbalanced tensegrity structure has laxity in its frame? (x) Why are obesity and menopause associated? (xi) What happens to the tensegrity after fasciotomies? (xii) If biology can really hide a pain condition in the tensegrity, what else has it hidden there? (xiii) Is having total belief in fibromyalgia a factor, does fear of death sustain it, and can one choose to willingly have fibromyalgia, even permanently?

Does “fascial armoring” fill in gaps regarding some of these anomalies? If affecting the trunk, a constrictive tissue in the chest might be able to impair lung function and contribute to wheezing and chest tightness. If involving a compression by the cervical fascia, it might affect salivary ducts/glands, affecting salivation. In the neck and pretracheal fascia, muscle tension dysphonia. Temporal fascia, tension like headache. Periorbital, reduced optic disc perfusion. Wrist, carpal tunnel syndrome. Hand, Raynaud’s-like phenomena. Under the skin, crawling sensations. Pelvic fascia, urinary urgency. Abdomen, peristalsis. Myofibroblasts contract over hours to lock in tension in ECM [51], so, under resting conditions stiffness might be exacerbated (e.g., sleep). Abnormal ECM properties could predispose to bruising. For impaired balance: proprioceptors are present among ECM, they affect coordination [18]. Sustained ECM forces might lead to their chronic activity, which, in turn, might activate reciprocal regions in the brain chronically. Chronic activity of those presynaptic nerve terminals in the brain might cause excitotoxicity, theoretically. A link to phenomena like seizures/pseudoseizures will be suggested below when mentioning the myodural bridge. Does central sensitization lead to small fiber neuropathy or does small fiber neuropathy lead to central sensitization? And how? According to this model, the answer is: neither-small fiber pathology in fibromyalgia abnormalities in Schwann cells seen on electron microscopy [200]. Neurofilaments provide mechanical strength and determine axon diameter, whereas microtubule functions include intracellular transport and provision of structural rigidity [201]. They were both found to exhibit appreciable changes in cytoskeletal spatial distributions within neuronal axons, under compressive loading [201]. Axonal changes under compressive loading are caliber dependent [201], neurite changes are dependent on substrate stiffness and gradient [202]. Substrate stiffness affects behavior and function of Schwann cells [203].

This theoretical model can only go as far as the cellular process that is behind it, so, what happens when modulating the cellular process? Interestingly, studies suggest certain antidepressants can downregulate myofibroblasts and ACTA2 gene expression [204,205], while other antidepressants may actually enhance them [206]. It seems gp120 has a stimulatory effect on myofibroblasts as well [71,207]. We could similarly predict that the pathogen Borrelia may lead to connective tissue changes and drive a widespread myofibroblast pathology. Thus, resulting in an infectious version of fibromyalgia with or without pronounced inflammation. Borrelia and fibroblast co-cultures show a significant induction of type I collagen mRNA after 2 days (p-value <0.02) and a significant upregulation of mRNA expression of TGF-β (p-value <0.01) [208]. Since TGF-β is a cardinal signal for myofibroblast α-SMA synthesis, this may explain why, of all infections, Lyme disease (and not viral encephalitis or infective transverse myelitis) is strongly associated with fibromyalgia. Due to the marked similarity to fibromyalgia, such events can be expected in connective tissue in “post COVID-19 syndrome”. Unudurthi et al. [209] suggest the patients can promote the activation of fibroblasts to myofibroblasts in heart. SARS-CoV-2 may induce the trans-differentiation of adipocytes or pulmonary lipofibroblasts into myofibroblasts, cells that play an integral part in fibrosis [210]. Since ECM remodeling is an ongoing chronic process, fibromyalgia-like symptoms would arguably be a sequela of infection rather than an acute manifestation.

#### Fascial armoring and the mind

A study suggested that in fibromyalgia, a global pathology involving tryptophan might come at the expense of other serotonergic operations such as in the brain [211]. A mechanosensitive effect can trigger serotonin release from enterochromaffin or neuroendocrine cells [212,213]. Serotonin is involved in the cellular signaling of connective tissue cells [214,215]. Studies of fibromyalgia patients indicate an elevated level of serotonin in myofascial tissue and platelets, while a decreased level of tryptophan and its derivatives, serotonin and kynurenine, is observed in the blood and/or brain [92,211,216–221]. Some fibromyalgia patients are found to have decreased rate of transport of tryptophan across the blood brain barrier [211]. Theoretically, a peripheral sequestration or depletion of biogenic amine metabolites may play a role in the association between mood disorders and fibromyalgia in some individuals and might explain why antidepressants seem to have such a limited efficacy in a subset of them.

Can an imbalanced tensegrity structure be associated with an imbalanced mind? It is possible some individuals feeling persistent tension and crawling sensations might try self-mutilation in such a way that physically releases tensegrity tension, rather than actually attempting suicide. Consciously or unconsciously: similar to pica, mentioned earlier. Associating pica with iron deficiency, glucosuria with diabetes, and self-harm with a tensegrity abnormality, seems to be a better choice for Nature than the other way around. Pica with “fascial armoring” would have pointed the author towards the mind. Lacerations would probably be the best first approach for this purpose and horizontal incisions might be more effective if vertical to force vectors. Unfolding this reason while using the scientific terms would be difficult for most common individuals, even for those with an extremely high body awareness; it would probably be described as: “trying to release a feeling”.

#### Compression versus a global percutaneous needle fasciotomy: conclusion

Studies show obesity induced connective tissue fibrosis is dependent on mechanosensitive signaling [222]. Fibrosis, via myofibroblasts, alters subcutaneous tissue plasticity and increases connective tissue rigidity and stiffness [222]. Thus, obesity might be associated with fibromyalgia by way of mechanically compressing fascia with higher forces. Is there any evolutionary sense? Unfortunately, obesity is one of the most “punished” conditions by nature. Does insulin-like growth factor affect central sensitization, and is weight gain from pregabalin helpful or harmful? A study in vivo suggests applying tension on skin induces myofibroblasts [74]. Therefore, by this same rationale we might expect a sedentary and/or hypermobile individual that tends to wear tight clothes and accessories that mechanically compress fascia to be at higher risk for this pathology over time. It was shown the association between a higher incidence of fibromyalgia and estrogen is unlikely [162,223]. Estrogen is suggested to inhibit myofibroblast differentiation and is associated with lower fascial stiffness [224–227], and hormonal contraceptives may reduce the risk of fibromyalgia in women [228]. Some suggest fibromyalgia may be linked to menopause [229]. This could be part of the explanation to why fibromyalgia is more prevalent in certain females. Other studies show that applying a splint or mechanical tension induces myofibroblasts [80], and ponytails/hijabs/braziers appear to cause myofascial pain [230–234]. This phenomenology appears to be similar to, or the same as, the old practice of applying bandages to treat “rheumatism” patients (William Balfour 1815 [235]), meaning: modulation of the tensegrity structure and/or induction of stress shielding but failing to resolve overall tension (e.g., the case of Mrs. M. pp. 179-180 [235]). Interestingly, experimental models of restraint stress apply mechanical restraint and/or limit movements [236–240]. These models seem to induce mechanical and cold allodynia, depression and anxiety like behavior, gut dysmotility, and other phenomena [236–240]. Thus, one might wonder what is the pluripotent entity that restraint stress tries to model? Is it only psychological stress?

In any case, perhaps a treatment or prophylaxis that applies multiple needles in different areas of the tensegrity system, could be utilized as a global percutaneous needle fasciotomy. The theoretical end purpose for this treatment would be by passively inserting the needles and then allowing the network of cells to lower global fascial stiffness beneath the threshold necessary for myofibroblasts activity (∼10-20 kPa) and slowly experience a tensional collapse. Each intrafascial (i.e., subcutaneous) needling therapy would have to be adapted according to the current state of the tensegrity structure, meaning: one protocol does not fit all, and does not fit one all times. If we ignore tensegrity principles, pain might increase. Patience seems key because the longer the duration of severe disease, the slower the change should be made. Human physiology does not tolerate sudden changes well, and “the entropy must go somewhere”. The myofibroblast that remain would be responsible for the relapse, at least in part. Elastography studies of fibromyalgia and MPS are scarce. Some studies found fascia of patients with myofascial pain measures statistically significantly higher on shear wave elastography, and they have increased peri-muscular connective tissue thickness and ultrasound echogenicity [241,242]. It was found that needling treatment reduces pain [243–247]. Studies suggest needling significantly lowers the shear modulus of myofascial tissue as measured by elastography (p-value <0.01) [95], causes mechanical changes at the point of insertion and at a distance [91,102], causes windup of the connective tissue [105], and improves perfusion to the area [248]. Meta-analyses and systematic reviews find that needling improves pain and stiffness in people with fibromyalgia and MPS [245,246,249]. Support for this modality’s capability of improving perfusion is found in a study showing infiltration of TrPs reduces symptoms of intermittent claudication [250]. Symptomatically, a person constantly exerting effort against such forces of a suffocating fascia would likely feel chronic fatigue and soreness, while accumulating nociceptive substances in muscles in a chronically tonic body, and active loci induced “energy crises”. Tryptophan usage would affect melatonin as well, and symptoms of cognitive impairment mean the brain is definitely involved. With regards to active loci, it is possible the cell has its own ways to oppose this abnormality. When synaptic neuromuscular activity is maintained over time, an aggregated form of amyloid beta is seen to interfere with Ach release ex vivo [251]. One adopting this biological ethos might want to examine if amyloid beta is linked to “fascial armoring”, because studies suggest amyloid beta, like the cytoskeletal protein tau, is affected by or compensates for, mechanical stress [252–256].

At least we might want to exclude an organic fascial cause before perceiving these cases as purely psycho-functional disorders, even if the full mechanism is still unclear. Is there enough empirical evidence to recognize MPS and fibromyalgia as honorable organic diseases, like any other? Internal medicine is used to entities that are either immune related, infectious, malignant, endocrine, metabolic, genetic, cardiovascular, neurologic, traumatic, or toxic, but not familiar with many mechanical ones. If medicine overlooked another system causing pain besides fascia, like integumentary or skeletal, it would probably lead to overlying on psychology to explain that pain. Maybe it is not some mechanism that has yet to be discovered or measured. Maybe it has been both discovered and measured yet overlooked and misinterpreted. It is difficult to explain all the empirical evidence relating to fibromyalgia (e.g., association with hypermobility syndrome, small fiber neuropathy, and the complete resolution soon after surgery) while relying on central sensitization and stress or somatization alone. Therefore, investigating a link between fibromyalgia and a peripheral mechanical process might benefit our understanding.

The enthusiast of the principle of Occam’s razor (i.e., minimize multiplication of entities) might consider seeing other functional entities that are known to be associated with pluripotent TrPs [2,4,12,43,257,258], as “MPS”/“fibromyalgia” that is localized to certain anatomical areas; coupled with the reciprocal central nervous system changes. Purely biologically, there really is only one entity (S1 Fig 2 grossly illustrates this overlap of conditions). “Kelley’s textbook of rheumatology” and others suggest functional somatic syndromes to be on a continuum of one entity with fibromyalgia [122,154,175]. Myofascial TrPs reproduce the pain pattern of fibromyalgia and are related to widespread mechanical hypersensitivity [259]. A study found that the topography of fibromyalgia tender points consists mostly of myofascial active TrPs (r=0.78, p-value <0.001) [260]. In addition, MPS and fibromyalgia are suggested to be two sides of the same coin [2,154,261]. Etymology does not necessarily reflect pathophysiology, but the term “fibromyalgia syndrome” basically carries the same meaning as “myofascial pain syndrome” but in Greek. The analysis based on this scoping review offers one mechanical aspect as a part of a common rheumapsychoneurological mechanism.

### Final note and future potential

Based on this review, it seems that fascia is an intelligent and sophisticated tissue network, at least like any other tissue network we know. It is shown myo/fibroblasts have gap junctions that can couple and directly communicate with each other (Xu et al. [262]) and with other types of cells (with heart [263], hair follicles [264], cancer cells [265], etc.) [51,266–270]. These can contract as a unit or in patterns [271,272], affecting the organ’s functionality [53,273–280]. If this theoretical model of myofibroblast-generated-tensegrity-tension is verified empirically, it might be worth further exploring the possibility that they can couple in vivo (directly, or indirectly through myocytes/glial cells) to neurons. Studies suggest the meninges might be coupled to the cerebral cortex to form one large network with myo/fibroblsts [281–284]. The myodural bridge connects the extracranial occipital fascia with the dura [285]. Myo/fibroblasts seem to form a body-wide cellular network; They can respond to mechanical cues and exhibit spontaneous calcium oscillations and synchronized contractions [62,270–272,279,286–288], and fibroblast themselves can induce contracture through actin beta and gamma pathways [289]. The psychological significance of such electrical-mechanical patterns is hypothesized not to be negligible. We could consider the hypothetical possibility that this dynamic body-wide mechanoelectrical network of cells and fibers has an overlooked interplay with the network of the human nervous system. This idea might disguise some of the pathophysiology of self-psychosomatics and potentially help us understand why a remarkable 17 percent of primary care patients are diagnosed with somatic symptom disorder (SSD) (defined in studies as at least four or more unexplained symptoms [290]), why tender point counts are predictors of CWP in somatising subjects [198], and why SSD is associated, or overlaps, with fibromyalgia. There are those who believe tensions in the body are closely linked to our emotions and personality (e.g., W. Reich and A. Lowen [291,292]). We know biology does not separate itself into different medical specialties like we do occupationally.

On a spectrum of “normality” we may all have “MPS” to some extent, technically speaking. It seems to be a question of how much we have and how much it affects our life. A tradeoff is still being made, no longer by biology but by humans. Not regarding the molecular design of fascia, but it is an ongoing tradeoff between not moving and moving. As it seems to be a natural condition worsened by an un-natural lifestyle, we may say living sedentarily misses our evolutionary target (i.e., a “biological sin”). It is interesting a study by Younger et al. finds patients with myofascial pain have changes in grey matter in the hippocampus, anterior insula, and cingulate cortex, among several areas [293]: Forgetting our evolutionary purpose seems to impair our memory, while cognitive detachment from our body-awareness seems to impair our cognition, and because biogenic amine metabolites are low, the above suggests a lifestyle with scarcity of movement will rarely make us blissful. Looking through the given titles of these entities, we might find what we most want to find where we least want to look. A body that cannot move with total freedom will hold a mind that is seldom totally free of dis-ease. This aligns with the notion that the body and the mind are accepted to be one being. Just as we can say music is a “collection of noises” or a “divine melody”, so may we say MPS is “pulled muscles” or a “fascinating disorder of the fascia”. Assembling these conclusions and concepts to creatively connect this medical puzzle is the easier part of this not so easy effort; The major challenge now being the research needed to verify or disprove them, and moving forward with evidence on this topic while moving ourselves…

## Limitations

The main limitation of this scoping review is that a sole researcher evaluated the studies. It is based on other empirical studies where some are in vitro studies, as mentioned in parts of the discussion. Being a scoping review, although a broad searching strategy was applied, not every topic was searched systematically. A systematic search was done for several main key phrases, in five databases. Systematically searching for relevant side topics (i.e., topics not defined in the key phrases) was not feasible, unfortunately. For side topics, an attempt to find a reliable source of information, like a textbook or study (or both), was the strategy to gather information. In such cases an attempt to cite more than one study was made. Worth noting, no on topic studies were found when searching “fibromyalgia myofibroblast” or “fibromyalgia alpha smooth muscle actin”. “Epigenetics and myofibroblasts” was not in the search. Kynurenine and the remaining 4% post-parathyroidectomy were not analyzed. The full cytokine profile and the topic of vitamin D in fibromyalgia was not discussed although a study finds vitamin D might play a role in myofibroblast attenuation or de-differentiation [294], and a meta-analysis suggests fibromyalgia is associated with a unique cytokine profile: TNF-α, IL-6, IL-8, IL-10 and eotaxin/CCL24 [295], all of those found to be implicated in myofibroblast cellular signaling [296–300].

## Data Availability

All data is available in the manuscript file

## Acknowledgements

Professor Pnina Ohanna Plaut for assisting with methodology and editing, Dr. Bronya Gorney for assisting with editing and support, Carrie Rodomar, librarian of University of Nicosia medical school, for assisting with methodology.

## Contributions

This work was done by the sole researcher and author of this manuscript including conception, organization, literature review, integration of information, findings analysis, manuscript preparation and writing.

## Conflict of interest

The author declares no conflict of interests.

## Funding

None

## Supporting information

**S1 File. Supplementary material**.

## Supplementary material for

### Methods

During the process of the review, certain topics rose that not were not sufficiently covered in the literature found through the systematic search. Therefore, further literature (through searches that were not systematic) was gathered for these topics. These topics were: myofibroblasts contractions and generation of tension, neurology of myofascial pain, myofascial pain and movement, fascial properties, searching pubmed for the term “unexplained”, etc.

Using “ovid” engine to search EMBASE yielded 0 results for the phrase “fascia tension pain” (whereas 127 items were found on PUBMED) therefore a broader search was performed in this database using only two word combinations: “fascia tension” (yielded 10 results) and “fascia pain” yielded 15 results (N=25 combined)

A search on COCHRANE was not done for the phrases “fascia tension pain” and “fascia stiffness pain” as the reviewer sought for controlled trials for myofascial pain syndrome, more than 150 items from COCHRANE oriented for MPS sufficed for this purpose.

Searches for “sympathetic activity induced by pain” and “Spinal mobilization sympathetic nervous system” was expanded for a search in all fields from inception only in databases where no results were found in title/abstract.

For “myofascial pain syndrome” the search was done on Cochrane but was limited to the title field only as it was too broad. This yielded 138 results.

Information gathered was summarized in excel document and a long word document containing key information from items. The word document evolved into the article, over time.

### Needling

To grasp the framework of needling, the reader may try and imagine a geodesic dome connected not by straight solid bars, but imagine it connected by extremely thin sheets of spandex/elastane. Each sheet can change its spring constant. Then imagine continuously applying external forces to this dome from different directions.

Insertion of needles in many “scattered” points while at rest will allow the system to realign appropriately and accordingly to the inherent internal forces or pullies [22], eg, the skeleton. Now the reader may imagine this dome only in the shape of a human, and it can move.

---

**S1 Fig 1.**
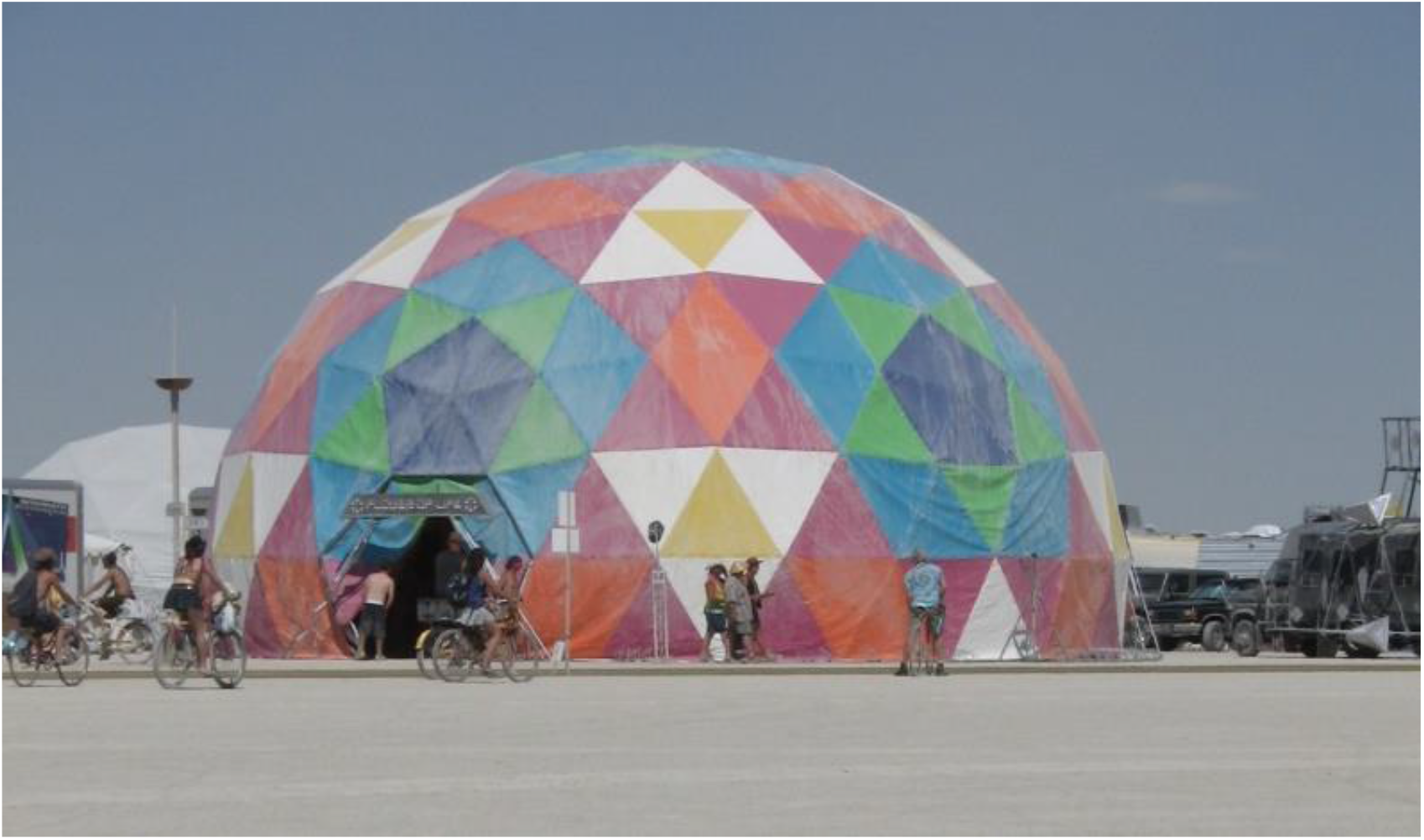
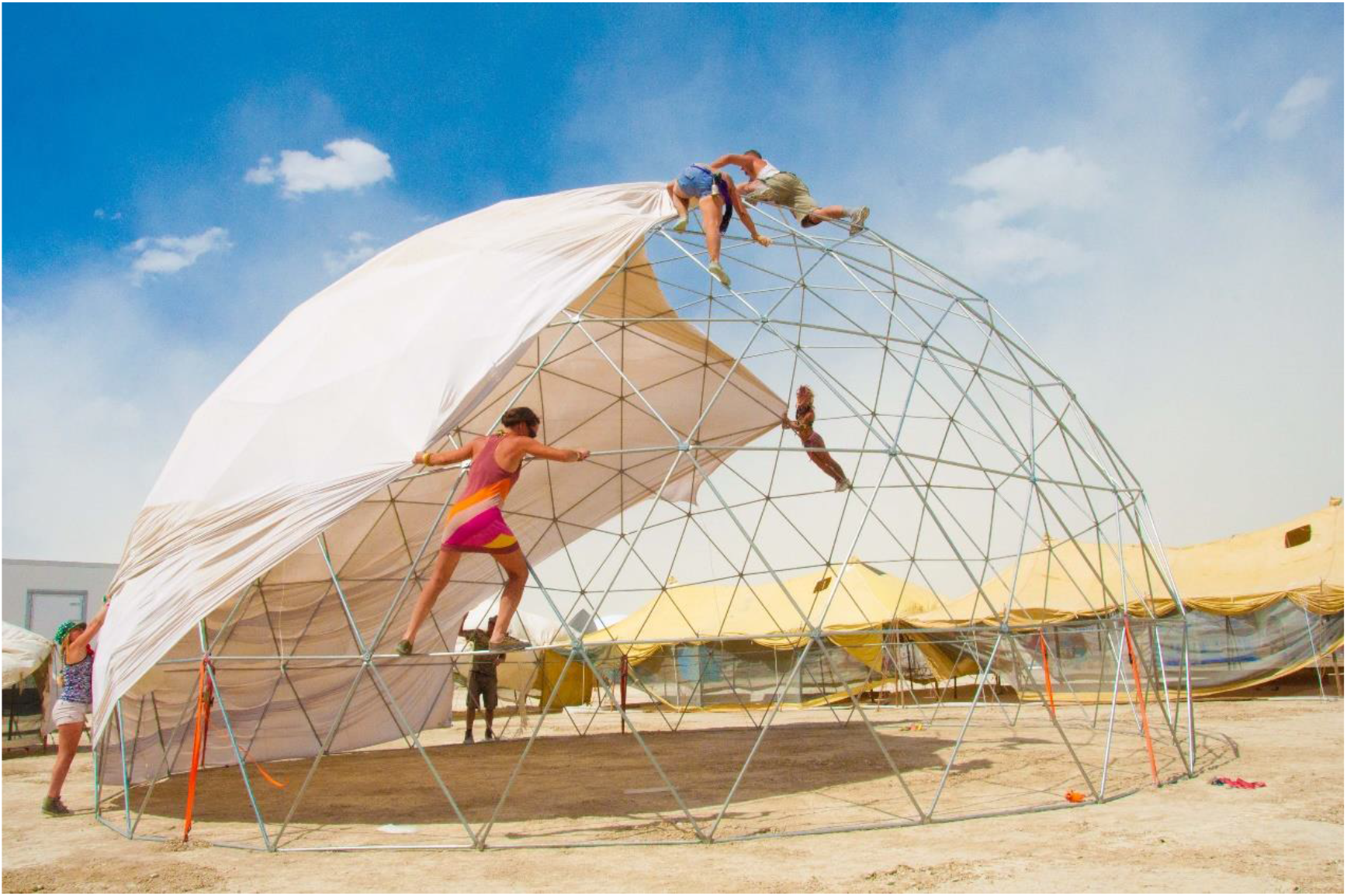
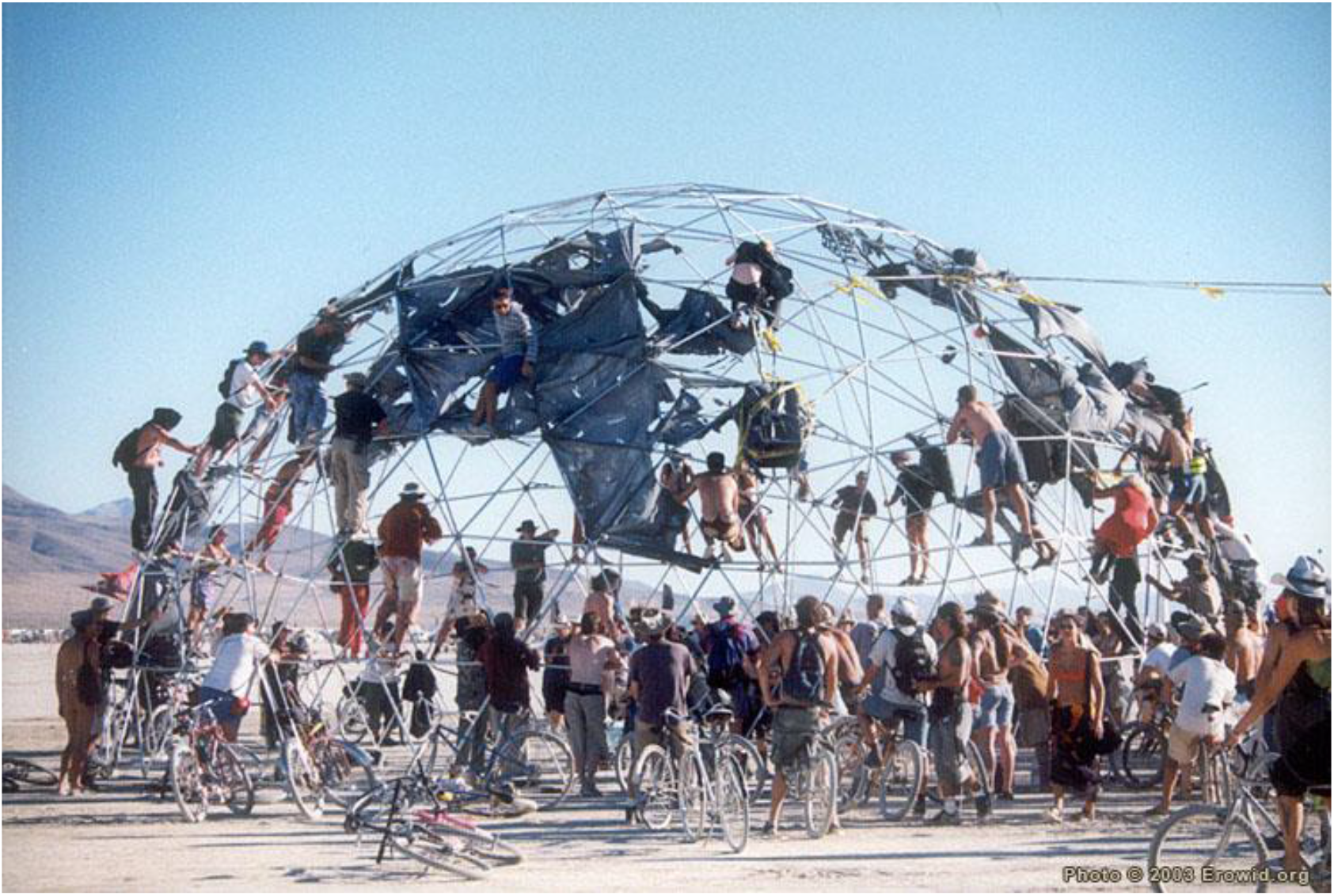
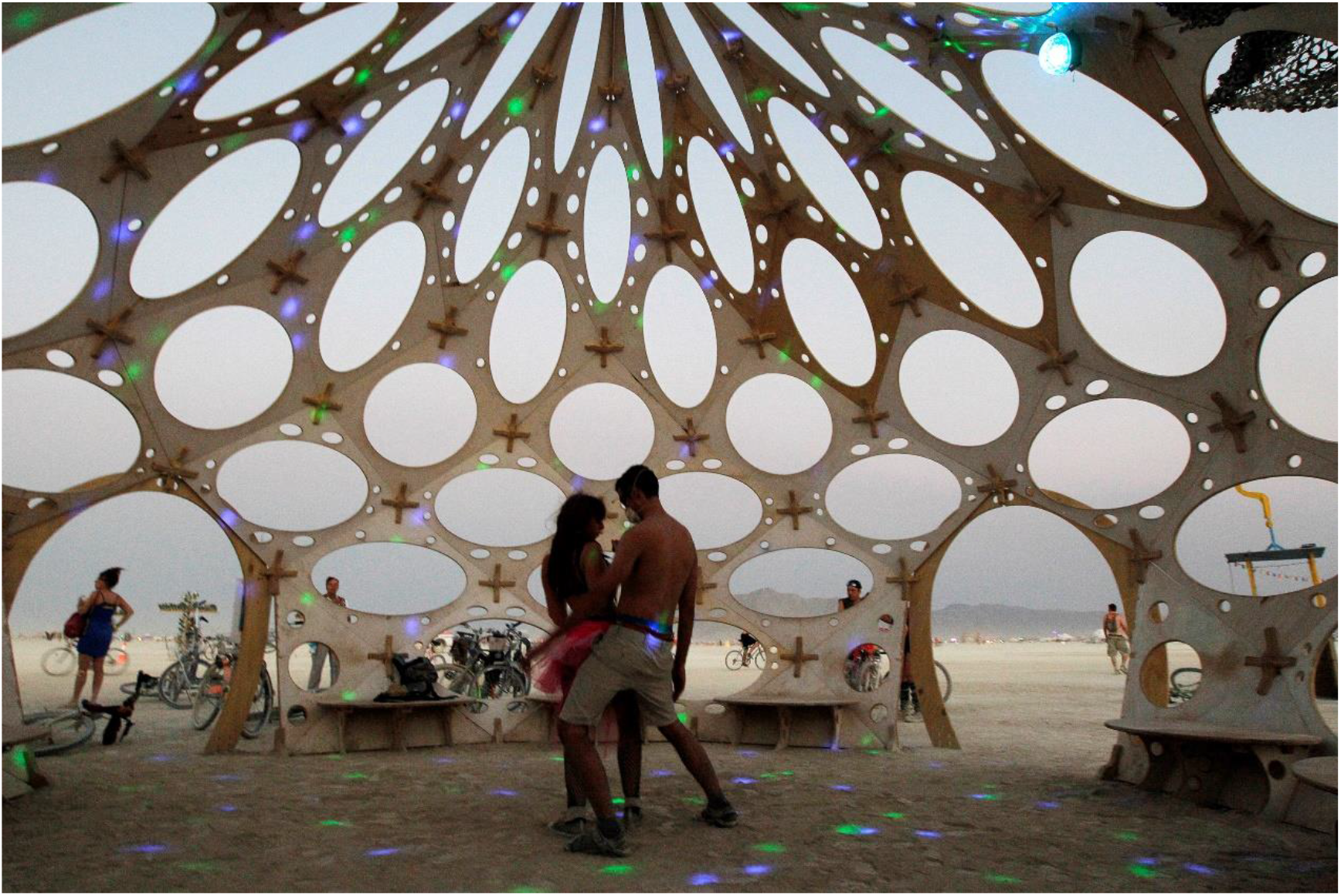
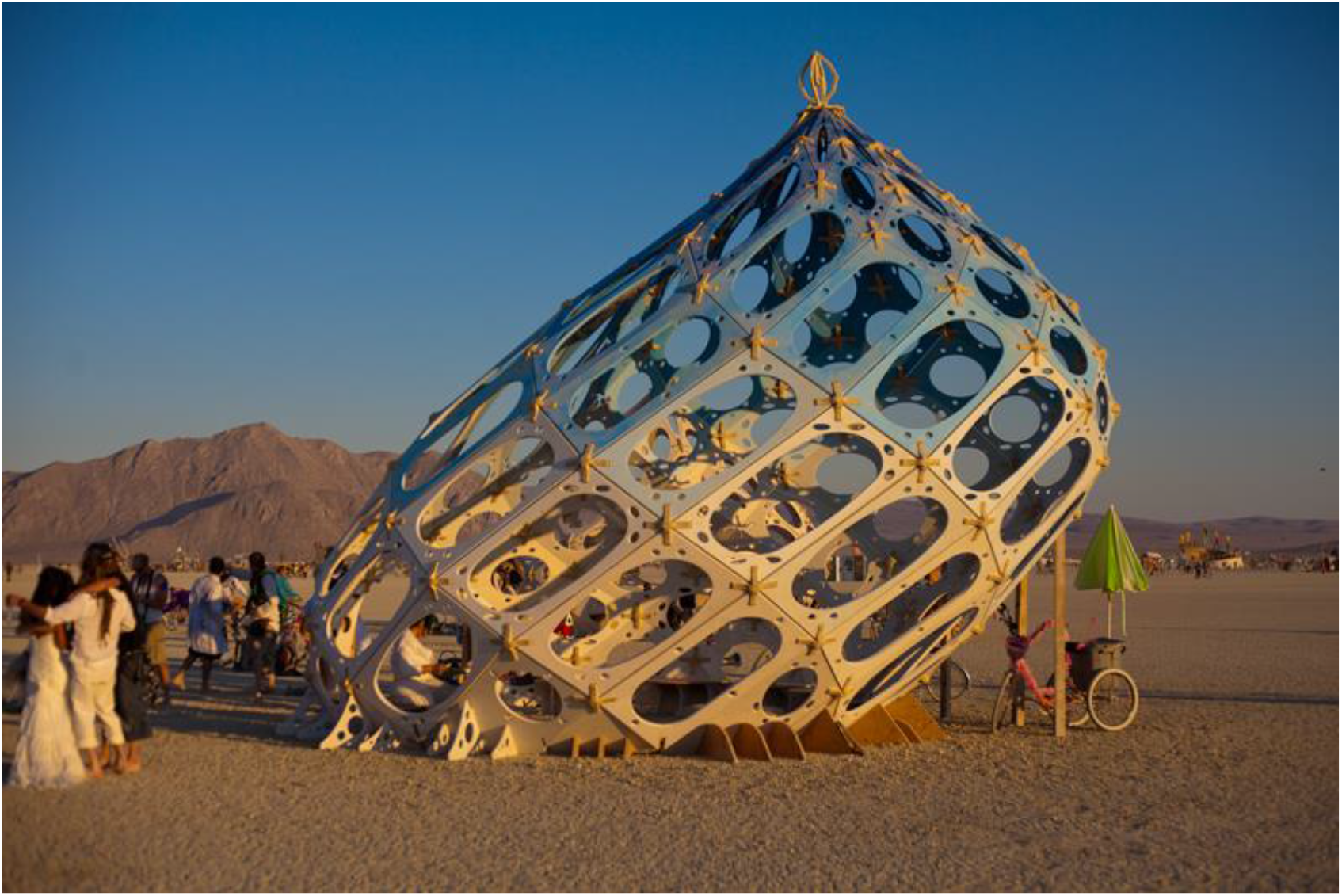
Illustration of needling in fascial tensegrity framework. A) Illustration to assist the reader in imagining a healthy geodesic dome connected by spandex sheets (fascia). With permission from PACIFIC DOMES Inc. www.eventdome.wordpress.com/ Reprinted from https://eventdome.files.wordpress.com/2010/07/bm-multi-colored.jpg, under a CC BY license, with permission from Sequoia Miller from Pacific Domes Inc., original copyright. B) Normal external strain applied will tense the fascia and distribute the tension through the network of nodes. With permission from Dome guys international www.domeguys.com Reprinted from https://domeguys.com/home/burning-man-2012-domeguys-international-kcj_-34/ under a CC BY license, with permission from Russell Phillips, President DomeGuys International LLC, original copyright 2021 C) Chronic over-strain causes pathological changes in fascia or changes of myofibroblast and smooth muscle fibers inside the fascia. The fascia fails to distribute forces properly and keep the integrity of the structure. Structures on the dome and inside the dome are affected. With permission from Erowid.org. Reprinted from https://www.erowid.org/culture/burningman/show_image.php?i=1999_burningman/1999_bm_thunderdome3.jpg under a CC BY license, with permission from original copyright 2021. D) After ‘re-setting’ the system with needling or other techniques: “regeneration and growth of new connections over time should be determined by natural forces and pullies” [22]. New areas in the fascia serve as points of focus for changes and realignment. Structures can relax and return to the minimum energy state. Fascia is always subjected to remodeling pressures and responds to the local mechanical state. However, if spatial deposition of fibers is altered with respect to physiological conditions, the rebuilding will be pathological [22] Mobilization encourages correct healing in order to avoid formation of fibrosis [22]. Taken by Jim Bourg, with permission from REUTERS News Agency. Reprinted from Jim Bourg photo published in https://www.theatlantic.com/photo/2013/09/photos-of-burning-man-2013/100584/#img06, with permission from Marc Glanville REUTERS News Agency, original copyright 2021. E) A pathological entity in the form of a tensegrity abnormality, it is both systemic and asymmetrical or seemingly “unilateral”. With permission from, and taken by, Aaron Neilson-Belman AaronNeilsonBelman.com Reprinted from https://hippievanman.com/preview/burning-man-2011/ under a CC BY license, with permission from Aaron Neilson-Belman, original copyright 2013-2020

**S1 Fig 2.**
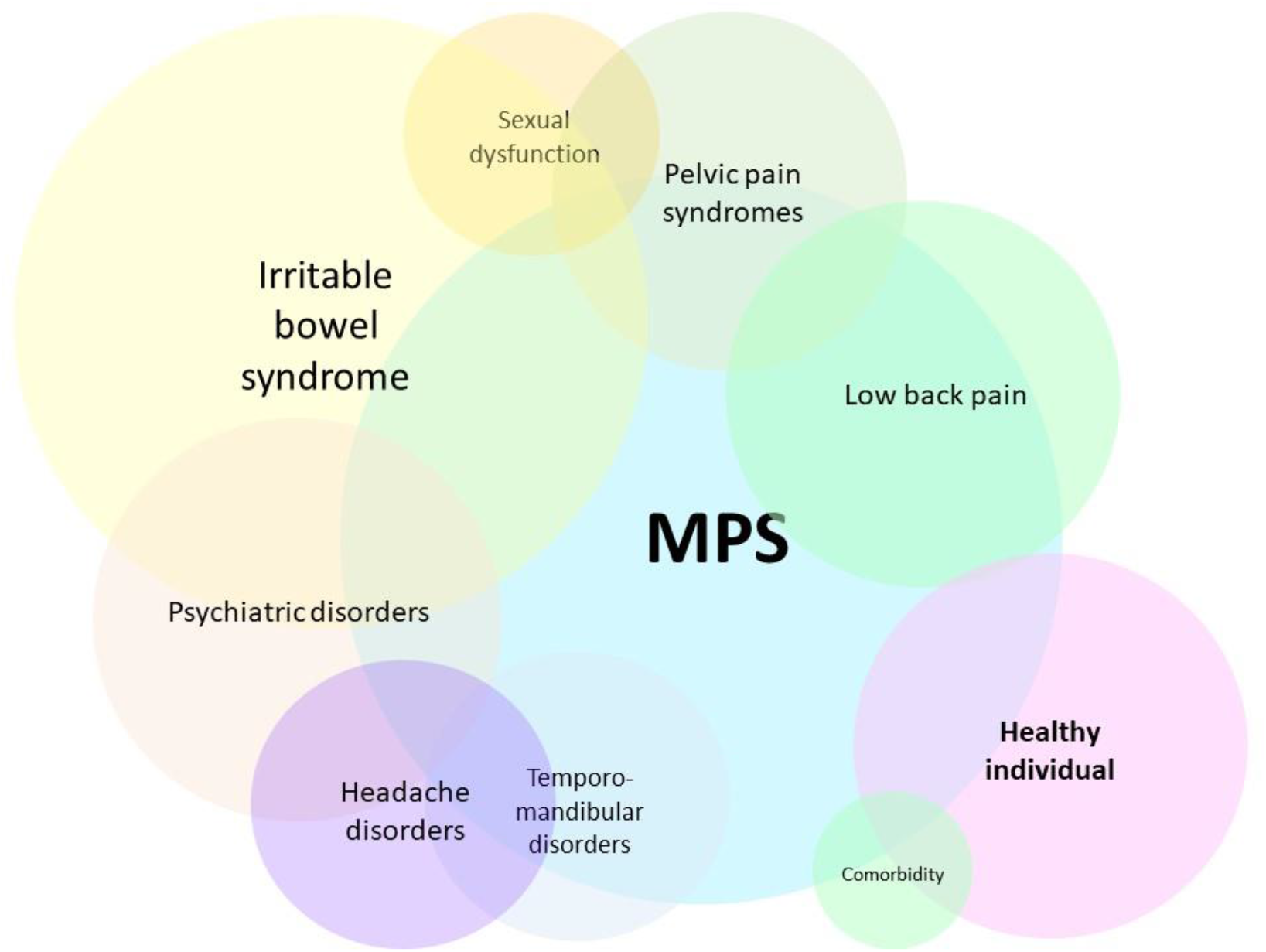
Overlap of conditions in the domain of common medicine. Not all connections are represented in this scheme. The term “Healthy” is open to interpretation. MPS-myofascial pain syndrome (i.e., fascial armoring).

## Notes

### Competing Interest Statement

The authors have declared no competing interest.

### Funding Statement

no funding

### Author Declarations

irrelevant to this manuscript

